# A Distinct Delayed-Onset Intraocular Inflammation Phenotype of Faricimab versus Aflibercept and Ranibizumab: Triangulated Pharmacovigilance Evidence from Databases of US, Europe and Japan

**DOI:** 10.64898/2026.10.10.26365218

**Authors:** Junhong Jiang, Yifan Gu, Di Hu, Xinsheng Li, Shiyue Qin, Qipeng Ling, Qi Zhang, Lili Dong, Zenan Lin

## Abstract

**Purpose:** Post-marketing reports have associated faricimab, the first intravitreal bispecific antibody, with intraocular inflammation (IOI). Its inflammatory phenotype, temporal pattern, and systemic safety relative to established anti-VEGF agents remain unclear. We compared faricimab with aflibercept and ranibizumab across three independent pharmacovigilance databases.

**Design:** Triangulated Pharmacovigilance study.

**Methods:** We analyzed FAERS reports (Q1 2022-Q2 2026) with external validation in JADER (Japan) and EudraVigilance (Europe). After de-duplication and restriction to primary-suspect reports, we computed reporting odds ratios (RORs) with false discovery rate control, derived confounder-adjusted RORs by logistic regression, modeled time-to-onset (TTO) with Weibull distributions, and graded signals with a pre-specified bidirectional cross-database evidence framework.

**Results:** The FAERS cohort comprised 6,833 faricimab, 6,714 aflibercept, and 2,533 ranibizumab reports. Faricimab showed higher reporting of anterior-chamber and vitreal inflammation: adjusted RORs (comparator versus faricimab; values <1 indicate faricimab excess) were 0.27 (95% CI 0.18-0.40) for iridocyclitis and 0.45 (0.35-0.58) for uveitis versus aflibercept, and 0.13 (0.06-0.24) and 0.12 (0.06-0.20) versus ranibizumab. All four core IOI preferred terms were Confirmed in JADER against both comparators. Faricimab IOI arose late (median TTO 100 vs 7 days; *P* = 2.6E-14) with a markedly flatter Weibull distribution (shape 0.81 vs ≈0.42). Aflibercept predominated for endophthalmitis-spectrum and intraocular pressure events; ranibizumab for vitreous hemorrhage (confirmed in JADER). Faricimab showed lower reporting across cardiac, nervous system, vascular, and renal organ classes, confirmed in EudraVigilance. An unadjusted death signal did not survive adjustment and was absent in outcome-based JADER analyses.

**Conclusions:** Faricimab exhibits a reproducible, delayed-onset, anterior-predominant IOI phenotype alongside a comparatively favorable systemic safety profile. Clinicians should recognize that faricimab-associated IOI may arise weeks to months after injection.

**Highlights:** Faricimab showed higher reporting of anterior and intermediate intraocular inflammation than aflibercept and ranibizumab across three independent databases.

Faricimab-associated inflammation arose late, with a median onset of 100 days versus 7 days for the comparators.

Aflibercept predominated for endophthalmitis-spectrum events and ranibizumab for vitreous haemorrhage.

No drug showed excess death reporting after adjustment and outcome-field verification.

## 1. Introduction

Retinal and choroidal vascular diseases, including neovascular age-related macular degeneration (nAMD), diabetic retinopathy (DR) and retinal vein occlusion (RVO), are a major cause of vision loss worldwide[1]. Vascular endothelial growth factor (VEGF) is central to the pathogenesis of ocular neovascularization and retinal exudation[2, 3]. After the off-label ophthalmic use of bevacizumab[4], ranibizumab was approved in 2006[5] and aflibercept in 2011[6]. Intravitreal anti-VEGF therapy is now first-line treatment for these conditions[7]. Alongside their efficacy, ocular adverse events have been documented across the class, including intraocular inflammation (IOI), endophthalmitis, intraocular pressure (IOP) elevation, vitreous haemorrhage and retinal vascular occlusion. Systemic events, including cardiovascular, cerebrovascular and renal events, have also been reported[8–11].

Faricimab (faricimab-svoa; Vabysmo™) is the first bispecific monoclonal antibody approved for intraocular use[12]. It neutralizes both VEGF-A and angiopoietin-2 (Ang-2), a mediator of vascular destabilization and inflammation. Pivotal trials showed sustained efficacy in nAMD, diabetic macular oedema and RVO[13–15]. Post-marketing experience, however, has raised a specific concern. Several case series describe sterile IOI after faricimab[16, 17], often arising weeks to months after injection and occasionally after several uneventful administrations. Three questions remain unanswered by single-database analyses: whether such inflammation is quantitatively more frequent with faricimab than with established agents, whether it differs phenotypically and temporally, and whether the engineered Fc region and high molecular weight of faricimab translate into lower systemic reporting.

Existing pharmacovigilance studies of faricimab have methodological limitations. Most rely on one or two spontaneous reporting system[18, 19]. Few adjust for demographic confounders, correct for multiple testing, or examine masking bias and differential market duration. Many report “top signals” dominated by preferred terms (PTs) that describe indications, product issues or medication errors rather than adverse clinical events. Single-database signals are also vulnerable to notoriety bias. This matters for faricimab, whose IOI has received substantial publicity. Independent replication is therefore essential before causal or clinical inferences are drawn.

We conducted a triangulated pharmacovigilance study comparing faricimab with aflibercept and ranibizumab in three independent spontaneous reporting systems: FAERS (United States), JADER (Japan) and EudraVigilance (Europe). The design included five pre-specified features: (i) a priori exclusion of non-AE PTs; (ii) confounder-adjusted head-to-head reporting odds ratios; (iii) time-to-onset analysis with Weibull shape parameter classification; (iv) sensitivity analyses for masking and temporal alignment; and (v) a bidirectional cross-database evidence classification framework. We aimed to characterize the comparative ocular inflammatory phenotype and its temporal signature, the systemic safety profile, and the robustness of fatal-outcome signals for faricimab.

## 2. Methods

### 2.1 Data sources

As shown in Figure 1., this retrospective pharmacovigilance study used three independent spontaneous reporting systems. The primary analysis drew on the FDA Adverse Event Reporting System (FAERS), a publicly available post-marketing database that collects adverse event reports from healthcare professionals, manufacturers and patients. We extracted all FAERS quarterly data from Q1 2004 through Q2 2026. The analysis window was restricted to Q1 2022 onwards, the approval quarter of faricimab, for the primary head-to-head analyses, with the full database used as the background reference set. External validation was performed in the Japanese Adverse Drug Event Report (JADER) database (PMDA, all available years)[20] and in EudraVigilance (European Medicines Agency)[21]. EudraVigilance data were aggregate System Organ Class (SOC)-level case counts, retrieved from the public dashboard on September 20, 2026. Faricimab was approved in Japan in March 2022, so JADER analyses covered the contemporaneous post-approval period for all three drugs.

**Figure 1.**
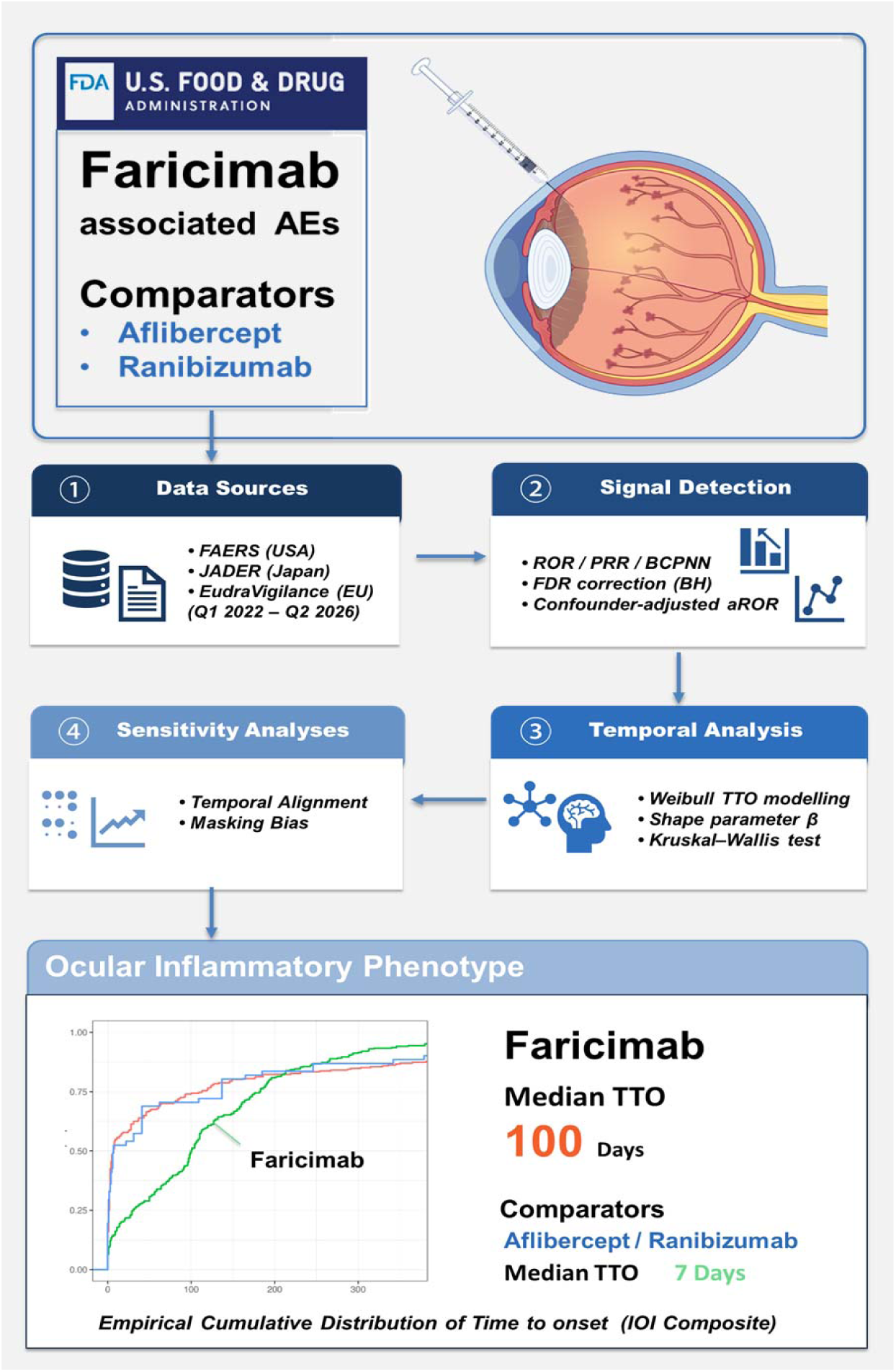
The Workflow.

Adverse events were coded to Medical Dictionary for Regulatory Activities (MedDRA) preferred terms (PTs) and mapped to SOCs using MedDRA version 27.1. Japanese PTs in JADER were translated to English with a targeted mapping table covering all pre-specified ocular and systemic events of interest. Unmapped PTs, which were predominantly non-ocular chemotherapy-related terms, remained in the denominators but could not contribute to event numerators (see 2.8).

### 2.2 Study drugs and cohort definition

Reports were eligible if faricimab, aflibercept or ranibizumab was recorded as the primary suspect (PS) drug. Duplicates were removed following FDA recommendations. For reports sharing the same CASE number, only the version with the most recent FDA receipt date was retained, with the highest case version number used to break ties. Reports listing more than one of the three study drugs in any role were excluded, ensuring mutually exclusive cohorts. To mitigate class-level masking, the background reference set excluded all reports mentioning any intravitreal anti-VEGF agent (faricimab, aflibercept, ranibizumab, brolucizumab or bevacizumab) in any drug role. A sensitivity analysis using the unmasked full database was also performed (see 2.7).

### 2.3 A priori exclusion of non-adverse-event preferred terms

Spontaneous reports for intraocular therapies frequently contain PTs that do not describe adverse clinical events. These include indication terms (for example “diabetic macular oedema”), product-quality and medication-error terms (for example “product issue” or “dose omission”), procedural terms (for example “vitrectomy”), terms describing absence of effect (“drug ineffective”), and ancillary investigation findings without clinical correlate. Such terms inflate both event counts and apparent signal strength. We therefore excluded a pre-specified list of 36 non-AE PTs (See Table S1) from all event-level analyses. Importantly, reports that contributed no remaining PT after this exclusion were retained in the denominators. RORs are thus computed against the full report population of each drug. Restricting denominators to reports with at least one retained PT would bias RORs upward.

### 2.4 Disproportionality analysis

For each drug–event pair we constructed 2×2 contingency tables and calculated the reporting odds ratio (ROR) with 95% confidence intervals[22], the proportional reporting ratio (PRR) with the chi-squared statistic (Yates-corrected)[23], and the Bayesian confidence propagation neural network (BCPNN) information component (IC) with its 2.5th percentile (IC025) using the Norén shrinkage estimator[24, 25].(See Table S2) A Haldane–Anscombe correction (0.5 added to each cell) was applied for zero cells. Analyses were performed at both SOC and PT levels. A positive signal required all of the following: at least 3 reports, lower bound of the ROR 95% CI above 1, PRR ≥2 with χ² ≥4, and IC025 >0. To control the false discovery rate across the large number of drug–event pairs, Benjamini–Hochberg adjustment was applied within each analysis family. Because unadjusted P values for the strongest signals underflow to zero in floating-point arithmetic, the adjustment was performed on the log10(P) scale using the rank-equivalent Benjamini–Hochberg formulation.

### 2.5 Head-to-head comparisons and confounder-adjusted RORs

Direct drug-to-drug comparisons were performed by computing RORs with each comparator’s reports serving as the other’s background. The full report cohort of each drug was used as the denominator. To account for differences in case-mix, we fitted multivariable logistic regression models to estimate adjusted RORs (aRORs) for pre-specified ocular and systemic events, with faricimab as the reference. Covariates were age group, sex, reporter region, reporter type, reporting year and indication; covariates with insufficient data for a given model were dropped automatically. Unadjusted and adjusted estimates are reported side by side.

### 2.6 Time-to-onset and Weibull shape parameter analysis

Time to onset (TTO) was defined as the interval between the therapy start date (THER table) and the event onset date (DEMO table), restricted to 0–1825 days. Events reported on the day of injection (TTO = 0) are common after intravitreal procedures; following established practice for Weibull modelling of spontaneous-report TTO data, these were assigned a value of 0.5 days[26]. For each drug and pre-specified event group (all AEs; an IOI composite of 12 PTs; ocular haemorrhage; arterial thromboembolism; death), the earliest event per case was retained. A Weibull distribution was fitted by maximum likelihood when at least 30 observations were available. The Weibull shape parameter β was classified as early failure (upper 95% CI bound <1), random failure (95% CI including 1) or wear-out failure (lower 95% CI bound >1). Between-drug TTO distributions were compared with the Kruskal–Wallis test.

### 2.7 Sensitivity analyses

Two sensitivity analyses addressed temporal and structural biases. First, faricimab entered the market in 2022, whereas ranibizumab and aflibercept have been marketed since 2006 and 2011. We therefore repeated the head-to-head comparisons after restricting all drugs to the contemporaneous post-2022 window. We also compared each comparator’s first three post-approval years with faricimab’s first three years to assess vulnerability to notoriety and Weber effects. Second, to evaluate masking, signals were re-computed against the full unmasked database. The correlation between masked and unmasked estimates was quantified with Spearman’s ρ.

### 2.8 External validation in JADER and EudraVigilance

In JADER, drugs were identified by Japanese generic and brand names (ファリシマブ/バビスモ; アフリベルセプト/アイリーア; ラニビズマブ/ルセンティス) and restricted to suspect-drug reports. The background was the full non-anti-VEGF database. Denominators comprised all reports of each drug and all background reports, not only reports containing a translated PT. This avoids inflating RORs when mapping coverage is incomplete. Death was ascertained from the structured outcome field rather than the PT “death”, because the two ascertainments diverge in spontaneous reports; the outcome-based analysis was pre-specified as primary. EudraVigilance provides aggregate SOC-level counts without case-level data. Head-to-head RORs were therefore computed from manually extracted SOC case counts for each drug.

### 2.9 Evidence classification framework

Each signal was classified into a four-tier evidence grade integrating direction and significance across databases. Head-to-head comparisons are directional: an ROR below 1 for “drug X versus faricimab” indicates higher reporting with faricimab. Classification was therefore performed bidirectionally. For each comparison and event, the direction of the signal was determined when the 95% CI excluded 1 and the higher side had at least 3 reports. Signals were graded as Confirmed (FAERS signal with at least one external database significant in the same direction), Consistent (external databases in the same direction by point estimate but not significant), Unconfirmed (FAERS signal without external replication), or Discordant (an external database significant in the opposite direction). Death was additionally evaluated using outcome-field ascertainment in JADER. Where PT-level and outcome-level results conflicted, the outcome-based result was considered definitive.

All analyses were performed in R version 4.4.2 using data.table, dplyr (1.2.1), fitdistrplus (1.2-6) and openEBGM (0.9.1). Reporting follows the READUS-PV guideline[27].

## 3. Results

### 3.1 Study cohorts

After de-duplication and restriction to primary-suspect reports in the contemporaneous window, the FAERS analysis cohort comprised 6,833 faricimab, 6,714 aflibercept and 2,533 ranibizumab reports (total N = 16,080), against a non-anti-VEGF background of 6,409,885 reports (See Figure 2.). The JADER validation cohort comprised 303 faricimab, 1,872 aflibercept and 1,156 ranibizumab suspect-drug reports, against a background of 1,024,966 reports. EudraVigilance contributed aggregate SOC-level counts only; case-level baseline characteristics are therefore not available for this source. Baseline characteristics of the FAERS and JADER cohorts are summarised in Supplementary Table S3.

**Figure 2.**
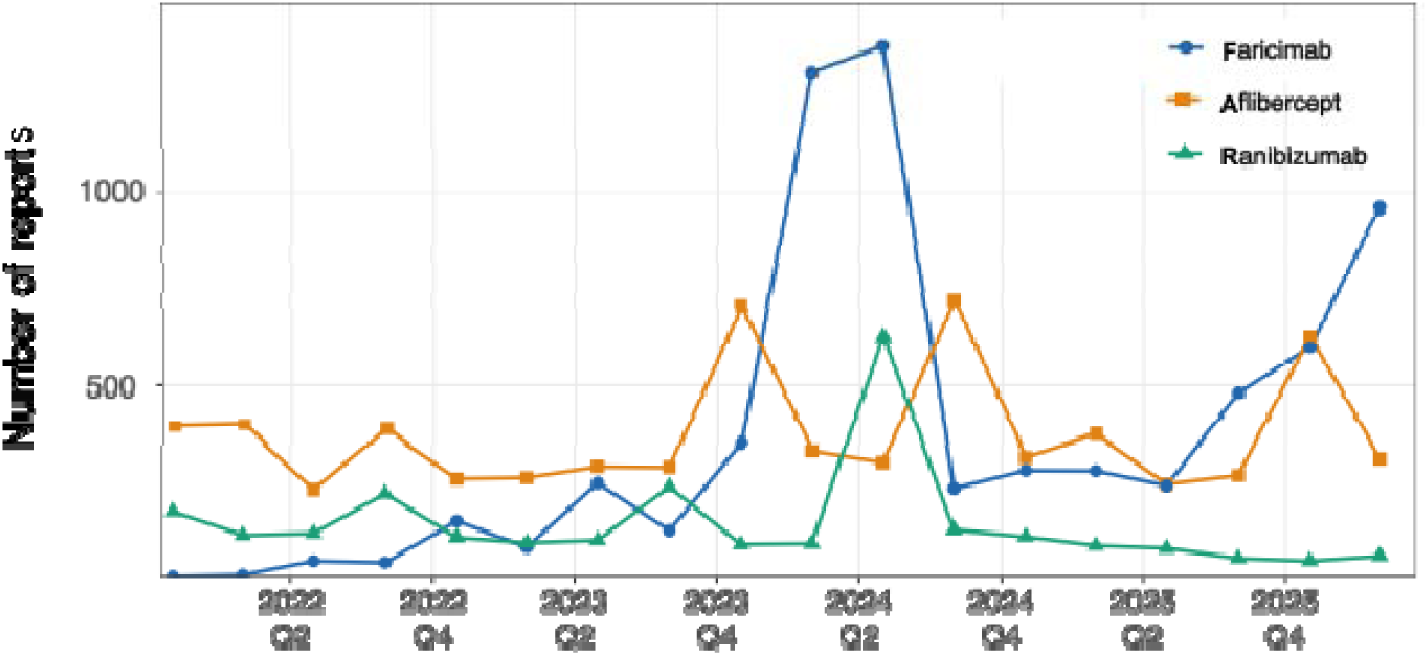
Quarterly trends of adverse event reports for intravitreal anti-VEGF agents in FAERS. Number of reports listing faricimab, aflibercept or ranibizumab as the primary suspect drug, by calendar quarter, from each drug’s first appearance in FAERS through 2026Q2. Quarters with no reports are shown as zero.

### 3.2 Disproportionality signals for faricimab versus the full database

After exclusion of the 36 pre-specified non-AE PTs, faricimab showed positive disproportionality for 70 PTs, dominated by ocular inflammatory and haemorrhagic events (See Figure S1. and Supplementary Table S4). At the SOC level, Eye disorders remained the dominant signal. Systemic SOCs including Cardiac, Nervous system and Vascular disorders showed negative disproportionality (ROR <1) (Supplementary Table S5).

### 3.3 Head-to-head comparisons: an intraocular inflammation phenotype split

Direct comparison revealed a consistent phenotypic split in the IOI spectrum (Table 1, Figure 3; full unadjusted and adjusted results in Supplementary Tables S6–S7, Figure S2–S3). Faricimab was reported significantly more often for anterior-chamber-predominant and vitreal inflammatory PTs than both comparators. For aflibercept versus faricimab, the adjusted RORs were 0.45 (95% CI 0.35–0.58) for uveitis and 0.27 (0.18–0.40) for iridocyclitis, where values below 1 indicate higher reporting with faricimab. For ranibizumab versus faricimab, the corresponding aRORs were 0.12 (0.06–0.20) and 0.13 (0.06–0.24). In the opposite direction, aflibercept was reported more often for endophthalmitis-spectrum and pressure-related events, including non-infectious endophthalmitis (ROR 5.25), anterior chamber cell (ROR 3.61) and intraocular pressure increased (aROR 2.61). Retinal vasculitis did not differ significantly after adjustment (aROR 0.89 versus aflibercept; 1.08 versus ranibizumab). Ranibizumab carried the highest reporting of vitreous haemorrhage (ROR 5.68, 95% CI 3.76–8.58 versus faricimab), a finding independently confirmed in JADER (see 3.6). Systemic SOC-level head-to-head estimates for all three pairwise comparisons are shown in Figure S4–S6.

**Figure 3.**
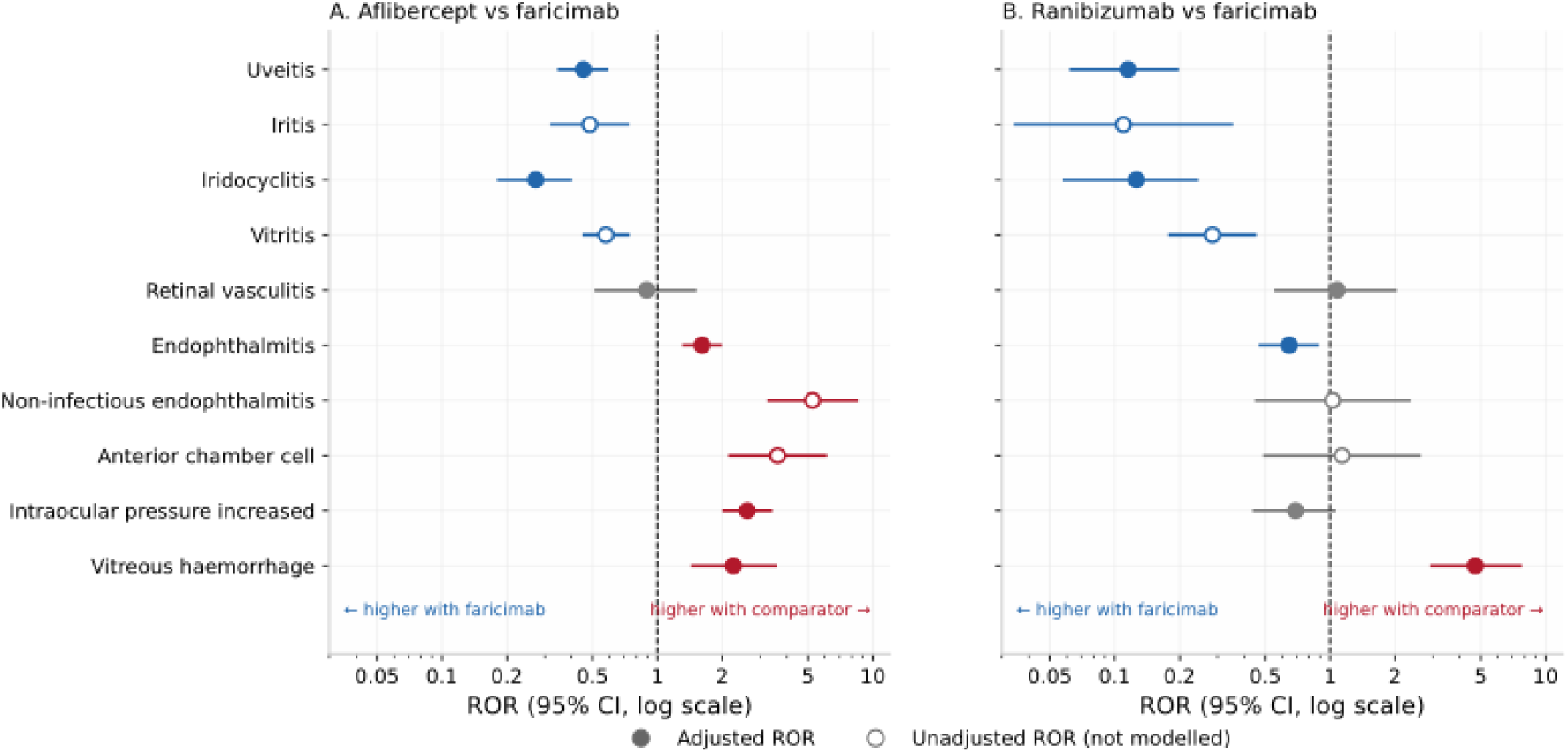
Head-to-head reporting odds ratios for the intraocular inflammation spectrum and selected ocular events. Forest plots of RORs (95% CI) for (A) aflibercept versus faricimab and (B) ranibizumab versus faricimab at the MedDRA Preferred Term level. Values below 1 indicate higher reporting with faricimab. Filled symbols denote covariate-adjusted RORs from multivariable logistic regression (adjusted for age group, sex, reporting year, region, reporter type and indication); open symbols denote unadjusted RORs for events that did not enter the adjusted models. The Haldane–Anscombe correction was applied for zero cells.

**Table 1.** Head-to-head comparison of the intraocular inflammation spectrum and selected ocular events: unadjusted and adjusted reporting odds ratios, FAERS (Q1 2022–Q2 2026).

| Preferred term |  | Aflibercept vs faricimab |  |  |  | Ranibizumab vs faricimab |  |  |  |
| --- | --- | --- | --- | --- | --- | --- | --- | --- | --- |
|  |  | *a* | *c* | ROR (95% CI) | aROR (95% CI) | *a* | *c* | ROR (95% CI) | aROR (95% CI) |
| Uveitis |  | 97 | 268 | 0.36 (0.28–0.45)* | 0.45 (0.35–0.58)* | 14 | 268 | 0.14 (0.08–0.23)* | 0.12 (0.06–0.20)* |
| Iritis |  | 35 | 73 | 0.49 (0.32–0.73)* | not modelled | 3 | 73 | 0.11 (0.03–0.35)* | not modelled |
| Iridocyclitis |  | 37 | 175 | 0.21 (0.15–0.30)* | 0.27 (0.18–0.40)* | 9 | 175 | 0.14 (0.07–0.27)* | 0.13 (0.06–0.24)* |
| Vitritis |  | 112 | 195 | 0.58 (0.46–0.73)* | not modelled | 21 | 195 | 0.28 (0.18–0.45)* | not modelled |
| Retinal vasculitis |  | 25 | 46 | 0.55 (0.34–0.90)* | 0.89 (0.52–1.49) | 17 | 46 | 1.00 (0.57–1.74) | 1.08 (0.56–2.01) |
| Endophthalmitis |  | 330 | 223 | 1.53 (1.29–1.82)* | 1.61 (1.33–1.96)* | 61 | 223 | 0.73 (0.55–0.97)* | 0.65 (0.47–0.87)* |
| Non-infectious endophthalmitis |  | 107 | 21 | 5.25 (3.29–8.40)* | not modelled | 8 | 21 | 1.03 (0.45–2.32) | not modelled |
| Anterior chamber cell |  | 67 | 19 | 3.61 (2.17–6.02)* | not modelled | 8 | 19 | 1.14 (0.50–2.60) | not modelled |
| Intraocular pressure increased |  | 226 | 110 | 2.13 (1.69–2.68)* | 2.61 (2.04–3.36)* | 30 | 110 | 0.73 (0.49–1.10) | 0.69 (0.44–1.05) |
| Vitreous haemorrhage |  | 65 | 34 | 1.95 (1.29–2.96)* | 2.25 (1.45–3.54)* | 70 | 34 | 5.68 (3.76–8.58)* | 4.72 (2.96–7.63)* |

*a* = number of comparator-drug reports listing the preferred term; *c* = number of faricimab reports listing the term. RORs below 1 indicate higher reporting with faricimab. ROR (95% CI) from 2×2 disproportionality analysis with the two drugs as mutual background (unadjusted). aROR (95% CI) from multivariable logistic regression with faricimab as reference, adjusting for age group, sex, reporting year, region, reporter type and indication; “not modelled” = the event was not among the pre-specified events entering the adjusted analysis. \* 95% CI excludes 1, with ≥3 reports on the higher-reporting side.

### 3.4 Time-to-onset and Weibull shape analysis

TTO analysis revealed a distinctive temporal pattern for faricimab-associated IOI (Supplementary Table S8, Figure 4). Median TTO for the IOI composite was 100 days (IQR 31–182) for faricimab, versus 7 days (IQR 2–113) for aflibercept and 7 days (IQR 1–137) for ranibizumab (Kruskal–Wallis *P* = 2.6×10□¹□). Only 6% of faricimab IOI events (24/377) occurred on the day of injection, compared with 16% (44/277) for aflibercept. Weibull shape parameters indicated early-failure patterns for all drugs and event groups (See Figure S7–S8). The faricimab IOI distribution, however, was markedly flatter (β = 0.81, 95% CI 0.75–0.88) than that of aflibercept (β = 0.41, 0.37–0.45) or ranibizumab (β = 0.43, 0.35–0.51). This is consistent with a delayed inflammatory phenotype rather than an immediate injection reaction.

**Figure 4.**
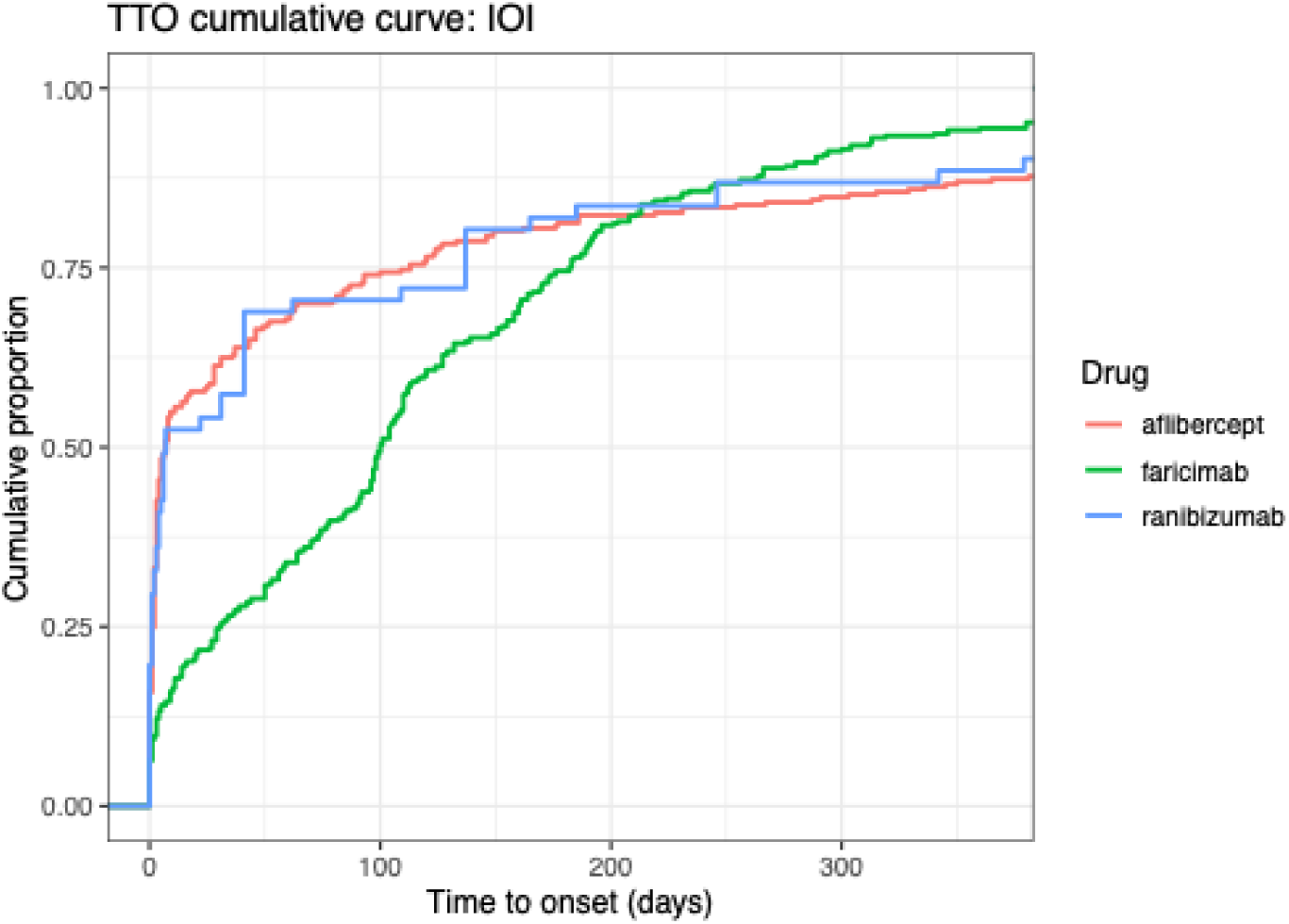
Time-to-onset distributions for the intraocular inflammation composite, by drug. Empirical cumulative distribution of time from therapy start to event onset (days) for the pre-specified IOI composite (12 PTs). Median TTO was 100 days (IQR 31–182) for faricimab versus 7 days for both comparators (Kruskal–Wallis P = 2.6×10□¹□). Weibull shape parameters were 0.81 (95% CI 0.75–0.88) for faricimab, 0.41 (0.37–0.45) for aflibercept and 0.43 (0.35–0.51) for ranibizumab.

Death showed a random-failure pattern for all three drugs, with every β 95% CI spanning 1. This argues against a time-clustered fatal toxicity. The median TTO of death was shorter for faricimab (188 days) than for aflibercept (561 days) (See Figure S9). This difference is expected under right-censoring: faricimab has been marketed only since 2022, so its fatal reports can only accumulate within a short window, whereas aflibercept reports span more than a decade. We therefore do not interpret this contrast as a pharmacological difference.

### 3.5 Sensitivity analyses

We also examined whether the choice of reporting window drove these results (See Figure S10 and Table S9). When each drug was restricted to its own first three post-approval years, Eye disorders was the only SOC that reached the signal threshold, and the magnitude was similar across drugs: ROR 12.89 (95% CI 11.48–14.47) for ranibizumab in 2006–2009, 18.03 (16.35–19.90) for aflibercept in 2011–2014, and 17.70 (16.67–18.78) for faricimab in 2022–2025. Heavy ocular reporting thus appears to come with the territory for a newly marketed intravitreal agent, rather than pointing to anything unusual about faricimab. If anything, faricimab compared favourably on systemic SOCs: its first-three-year RORs for Cardiac, Nervous system and Vascular disorders (0.30, 0.29 and 0.47) sat below the corresponding launch-era estimates for aflibercept (0.69, 0.43, 0.64) and ranibizumab (1.12, 1.08, 0.94). Notoriety bias, which inflates reporting in the first years after launch, would have pushed faricimab’s estimates upward; its systemic RORs were nonetheless the lowest of the three. Masking told much the same story. RORs computed against the full unmasked database tracked the masked estimates almost exactly (Spearman ρ = 0.984 at SOC level, 0.9996 at PT level; See Figure S11–S12, Supplementary Table S10), and no PT-level signal found under the masked background disappeared when the full database was used.

### 3.6 External validation in JADER and EudraVigilance

The faricimab IOI signal replicated in JADER for all four core PTs against both comparators (See Table S11a and S11b). For aflibercept versus faricimab, RORs were 0.048 (0.017–0.132) for iridocyclitis, 0.189 (0.092–0.388) for iritis, 0.243 (0.151–0.393) for uveitis and 0.196 (0.091–0.423) for vitritis. For ranibizumab versus faricimab, the corresponding RORs were 0.109 (0.045–0.268), 0.090 (0.032–0.251), 0.074 (0.035–0.158) and 0.063 (0.018–0.225). These values indicate roughly 4- to 20-fold higher relative reporting of these events with faricimab in both databases. Retinal vasculitis was directionally consistent but not uniformly significant (JADER ROR 0.55, 0.26–1.17 versus aflibercept; 0.199, 0.074–0.539 versus ranibizumab). In the opposite direction, vitreous haemorrhage was confirmed as a ranibizumab-predominant signal (JADER ROR 38.9, 95% CI 2.40–632.1 versus faricimab; 2.60, 1.73–3.91 versus aflibercept). Cerebral infarction was reported more frequently with both comparators than with faricimab.

Outcome-based death analyses in JADER showed no significant difference between any pair of drugs (See Table S11c). RORs were 0.84 (0.51–1.40) for aflibercept versus faricimab, 0.89 (0.53–1.51) for ranibizumab versus faricimab, and 1.06 (0.77–1.46) for ranibizumab versus aflibercept. The unadjusted FAERS death signal was therefore not confirmed.

At SOC level, EudraVigilance confirmed the systemic safety pattern (See Table S12). Compared with faricimab, aflibercept showed significantly higher reporting of Blood and lymphatic system, Cardiac, Hepatobiliary, Metabolism and nutrition, Nervous system, Psychiatric, and Renal and urinary disorders (See Figure S13). Ranibizumab showed significantly higher reporting of Blood and lymphatic system, Cardiac, Hepatobiliary, Nervous system, Psychiatric, Renal and urinary, and Vascular disorders (See Figure S14). In the ranibizumab versus aflibercept comparison, Nervous system and Vascular disorders were higher with ranibizumab.

### 3.7 Cross-database evidence classification

Applying the pre-specified framework (See Figure 5, Tables S13 and S14), the four core IOI PTs (uveitis, iritis, iridocyclitis, vitritis) achieved Confirmed status against both comparators, with FAERS significance independently replicated in JADER. Retinal vasculitis was graded Consistent. Endophthalmitis showed a split pattern: it was higher with aflibercept than faricimab, but higher with faricimab than ranibizumab. This suggests that the endophthalmitis signal of faricimab may be driven mainly by its non-infectious inflammatory component. Sixteen SOC-level signals of excess systemic reporting with the comparator drugs were Confirmed in EudraVigilance. The death signal was graded Discordant at PT level, but it was not supported by outcome-based ascertainment in JADER and disappeared after adjustment in FAERS. Visual impairment showed genuinely discordant directions between FAERS and JADER, plausibly reflecting national differences in how visual outcomes are reported.

**Figure 5.**
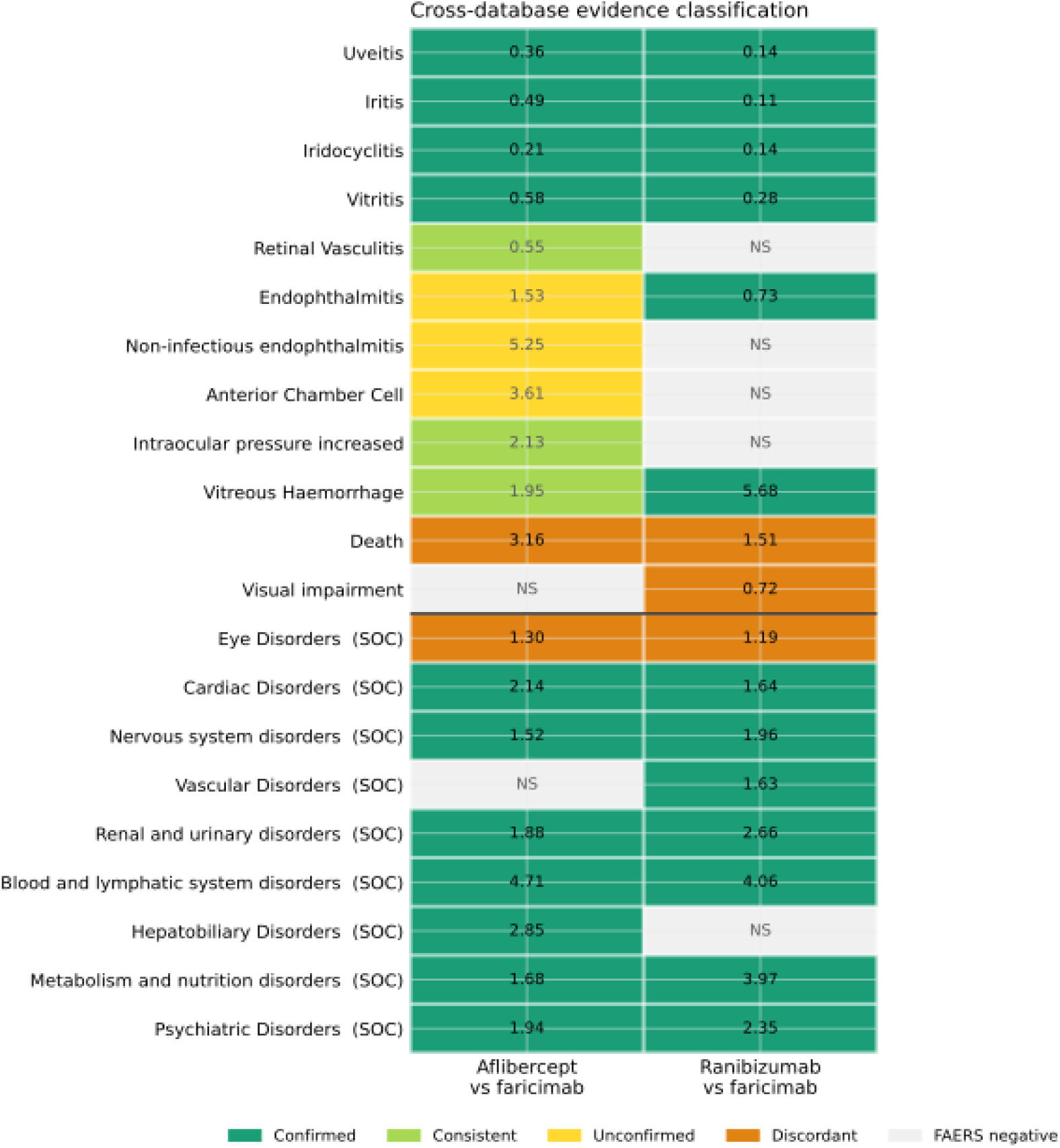
Cross-database evidence classification of pre-specified signals. Each cell shows the evidence grade for one signal in one head-to-head comparison, integrating direction and significance across FAERS with external validation in JADER (PT level) and EudraVigilance (SOC level). Signals were graded bidirectionally as Confirmed (FAERS signal replicated at significance in ≥1 external database), Consistent (same direction by point estimate without external significance), Unconfirmed (FAERS signal without external replication), Discordant (external significance in the opposite direction) or FAERS negative. Cell annotations show the FAERS ROR; “NS” indicates no FAERS signal. For death, the grade reflects PT-level ascertainment; outcome-field verification in JADER found no between-drug difference.

## 4. Discussion

### 4.1 Principal findings

In the largest head-to-head post-marketing comparison of faricimab, aflibercept and ranibizumab to date, we found a consistent and reproducible split in the intraocular inflammation phenotype across three independent spontaneous reporting systems. Faricimab was reported disproportionately often for anterior and intermediate uveitic events: iritis, iridocyclitis, uveitis and vitritis. The signal was significant in FAERS for both comparators and replicated at similar magnitude in JADER. EudraVigilance, at SOC granularity, pointed in the same direction. These events also carried a distinctive temporal signature. The median onset of 100 days, the flat Weibull distribution and the low proportion of same-day reactions all distinguish faricimab IOI from an acute injection reaction. In the opposite direction, aflibercept was reported more often for endophthalmitis-spectrum events, anterior chamber cell and increased intraocular pressure, while ranibizumab predominated for vitreous haemorrhage. After adjustment and outcome-field verification, no excess death reporting was seen for any of the three drugs.

### 4.2 The IOI phenotype split: possible mechanisms

Why would one intravitreal antibody shift the inflammatory phenotype towards anterior and intermediate segments? Several non-exclusive mechanisms deserve consideration. First, molecular structure differs fundamentally between the agents. Aflibercept is a soluble decoy receptor with high VEGF-A affinity[28]; ranibizumab is a Fab fragment lacking an Fc region[29]; faricimab is a full-length IgG with an engineered Fc that binds both VEGF-A and angiopoietin-2[30]. Fc-bearing molecules can engage Fcγ receptors on resident ocular immune cells, and immune-complex formation has long been implicated in uveitic reactions to systemically administered biologics. The brolucizumab experience showed that full-length constructs are not automatically inflammatory, but it also showed that small differences in molecular design can produce large differences in intraocular immunogenicity[31]. Second, Ang-2/Tie2 blockade is pharmacologically distinct from pure VEGF inhibition. Tie2 signalling modulates endothelial stability and pericyte function, and its inhibition could alter blood–ocular barrier permeability or the set-point of intraocular immune surveillance in ways that favour delayed, anteriorly distributed inflammation[32]. Third, the temporal pattern we observed is hard to reconcile with a toxic injection reaction. Acute post-injection inflammation typically peaks within days. The 100-day median onset and β close to 1 instead resemble an immune-mediated, possibly cumulative mechanism that unfolds over repeated dosing intervals, analogous to the delayed vasculitic pattern documented for brolucizumab[31]. Fourth, formulation factors cannot be excluded. Buffer composition, protein concentration and particle burden differ between products and may interact with the vitreous environment to determine where inflammation localises.

The reverse signals also merit mechanistic comment. The higher reporting of non-infectious endophthalmitis and anterior chamber cell with aflibercept has precedents in early post-approval clusters of sterile inflammation, some of which were linked to syringe silicone-oil interactions[33]. Ranibizumab’s vitreous haemorrhage predominance may partly reflect its older indication mix, with more proliferative diabetic retinopathy in its treated population, although the signal persisted in JADER after contemporaneous restriction[34]. Ranibizumab was also the only drug with a confirmed vascular SOC signal (See Table S14). This echoes the historical debate over systemic VEGF exposure from intravitreal Fab fragments[35]. Our design cannot settle causality, but the cross-database convergence strengthens the case that these are drug-level rather than database-level phenomena.

### 4.3 The death signal: a cautionary tale for spontaneous-report analysis

Unadjusted FAERS analysis suggested higher death reporting with faricimab (See Table S7). This signal did not survive scrutiny. Three independent lines of evidence argue against a true excess. First, the signal disappeared after adjustment for age, sex, region, reporter type, reporting year and indication. Second, outcome-based ascertainment in JADER found no between-drug difference in fatal outcomes, whereas PT-level death counts diverged, indicating an ascertainment artefact (See Table S11). Faricimab reports are more likely to carry the literal PT “death” without a corresponding fatal outcome field. Third, the Weibull analysis showed a random-failure pattern for death with all three drugs, with no temporal clustering (See Table S8). The shorter median TTO of faricimab death reports is an arithmetic consequence of its short market window, not a pharmacological property. Together these findings illustrate why unadjusted PT-level death signals in spontaneous reports should be treated as hypothesis-generating at best, and why outcome-field verification and adjustment are essential before any mortality claim is made.

### 4.4 Comparison with previous studies

Pivotal programme data showed comparable non-infectious IOI between faricimab and aflibercept in TENAYA/LUCERNE[36] and YOSEMITE/RHINE study[32]. Post-marketing case series have described faricimab-associated uveitis and retinal vasculitis often occurring after multiple injections and responding to corticosteroids[16, 37, 38]. Our results quantify these case-level observations at population scale and add two elements that case series cannot provide: comparator context and temporal structure. The finding that retinal vasculitis itself was not significantly elevated after adjustment, despite the strong anterior IOI signal, suggests that faricimab IOI is phenotypically distinct from the brolucizumab vasculitic syndrome, in which retinal vascular involvement was the dominant feature[31]. Whether a small subset of faricimab IOI cases progresses to occlusive vasculitis remains an important clinical question that spontaneous reports cannot answer.

### 4.5 Strengths

This study has several strengths. It is, to our knowledge, the first triangulated head-to-head pharmacovigilance analysis of faricimab across three regulatory jurisdictions, covering more than 19,000 drug-attributed reports in the primary and validation cohorts. The bidirectional evidence classification framework avoids a common pitfall of head-to-head disproportionality work, in which the direction of an ROR below 1 is misread and signals on the wrong side are counted as null. The pre-specified exclusion of non-AE PTs, and the denominator-preserving analysis design, protect against two further biases that are widespread in the published disproportionality literature. Adjustment for case-mix, outcome-field verification of death, masked-versus-unmasked sensitivity analyses and contemporaneous-era restriction all point the same way: the principal findings are robust.

### 4.6 Limitations

Several limitations must be acknowledged. First, spontaneous reports cannot establish incidence or causality. Reporting ratios are not risk ratios, and stimulated reporting around faricimab’s launch almost certainly inflated early IOI reporting, although the contemporaneous-era sensitivity analysis and the replication in JADER, where faricimab was approved in the same period, make pure notoriety bias an unlikely full explanation. Second, case-level clinical detail is limited. We could not verify injection counts, laterality, dose or prior anti-VEGF exposure, all of which influence IOI risk, and we could not distinguish infectious from non-infectious endophthalmitis beyond PT wording. Third, TTO analyses are right-censored for faricimab. Its short market window truncates the observable TTO distribution and biases median onset downwards for any event. The direction of this bias works against the delayed-onset finding, so the IOI temporal contrast is likely conservative, but it prevents any comparison of long-latency events. Fourth, JADER analyses relied on a targeted PT translation table. Coverage of ocular terms was high, yet residual unmapped PTs could attenuate event numerators while denominators were complete, biasing JADER RORs towards the null. Fifth, EudraVigilance contributed only aggregate SOC-level counts, precluding PT-level confirmation and covariate adjustment. Finally, although we adjusted for indication, indication mix differs between drugs and eras, making residual confounding by disease severity possible.

### 4.7 Clinical and regulatory implications

For clinicians, the practical message is not that faricimab is unsafe. The absolute reporting burden remains low relative to its global use, and no mortality signal was confirmed. Rather, patients receiving faricimab warrant specific counselling about delayed ocular inflammation, with attention to anterior segment symptoms arising weeks to months after injection, and a low threshold for slit-lamp review. Unilateral switching from aflibercept to faricimab in eyes with prior IOI should be considered cautiously until the cross-reactivity question is resolved. For regulators, the convergent tri-database signal supports continued inclusion of uveitic events in faricimab’s periodic safety reviews and suggests that label language distinguishing delayed anterior IOI from acute post-injection inflammation would aid recognition. For researchers, our framework demonstrates how bidirectional head-to-head comparisons, outcome-field verification and TTO characterisation can be combined to turn a raw disproportionality signal into graded, clinically interpretable evidence.

## 5. Conclusions

Across three independent pharmacovigilance databases, faricimab showed a reproducible, temporally distinctive signal of delayed anterior and intermediate intraocular inflammation relative to aflibercept and ranibizumab, without confirmed excess retinal vasculitis or death. Aflibercept and ranibizumab carried their own distinct profiles, dominated by endophthalmitis-spectrum events and ocular haemorrhage respectively, plus a heavier systemic SOC burden. These findings do not alter the overall benefit–risk balance of any agent, but they support phenotype-aware monitoring and informed counselling for patients receiving intravitreal anti-VEGF therapy.

## Data Availability

FAERS data are publicly available from the FDA website (https://www.fda.gov/drugs/drug-approvals-and-databases/fda-adverse-event-reporting-system-faers-database). JADER data are publicly available from the PMDA website (https://www.pmda.go.jp/safety/info-services/drugs/adr-info/suspected-adr/0003.html). EudraVigilance data were obtained from the public dashboard (https://www.adrreports.eu/en/search_subst.html). Analysis code is available on GitHub repository through https://github.com/DrWoodWood/faricimab-pv.

https://www.pmda.go.jp/safety/info-services/drugs/adr-info/suspected-adr/0003.html

https://www.adrreports.eu/en/search_subst.html

## Declarations

### Ethics approval and consent to participate

This study used publicly available, de-identified spontaneous reporting data (FAERS, JADER, EudraVigilance public dashboard). No institutional review board approval was needed.

### Competing interests

The authors declare no competing interests.

### Funding

This study was supported by the Taizhou Science and Technology Support Plan social development project grant (TS202418), the Natural Science Foundation of Jiangsu Province (BK20250567) and the National Natural Science Foundation of China (82401257).

### Authors’ contributions

J.J., Y.G. and D.H. downloaded and analysed the drug safety data; J.J., Y.G., D.H., X.L., S.Q., and Q.L. wrote the manuscript. Z.L. conceived and designed this study. Q.Z., L.D. and Z.L. supervised this work and reviewed the manuscript. All authors approved this work.

## Acknowledgements

The authors thank the researchers and administrators of FAERS, JADER and EudraVigilance for making the drug safety data publicly available.

## Tables

**Table 1.** Head-to-head disproportionality for pre-specified ocular and systemic events: unadjusted and adjusted RORs for aflibercept versus faricimab and ranibizumab versus faricimab.

**Supplementary Table S1.** Baseline characteristics of the FAERS and JADER analysis cohorts.

**Supplementary Table S2.** Preferred terms mapped to each System Organ Class.

**Supplementary Table S3.** The 36 pre-specified excluded non-adverse-event preferred terms.

**Supplementary Table S4.** SOC-level disproportionality for faricimab versus the masked full-database background.

**Supplementary Table S5.** PT-level disproportionality signals for faricimab (70 positive PTs).

**Supplementary Table S6.** Head-to-head SOC-level disproportionality (all three pairwise comparisons).

**Supplementary Table S7.** Head-to-head PT-level disproportionality (all three pairwise comparisons).

**Supplementary Table S8.** Time-to-onset and Weibull shape parameter analysis, by drug and event group.

**Supplementary Table S9.** Era-restricted sensitivity analyses: contemporaneous window and first-three-years comparison.

**Supplementary Table S10.** Masked versus unmasked background sensitivity analysis.

**Supplementary Table S11.** JADER head-to-head disproportionality (PT level).

**Supplementary Table S12.** EudraVigilance SOC-level head-to-head disproportionality.

**Supplementary Table S13.** Evidence classification framework definitions.

**Supplementary Table S14.** Cross-database evidence classification of all evaluated signals.

## Figure legends

**Figure 1**. Quarterly trend of spontaneous reports for faricimab, aflibercept and ranibizumab in FAERS, Q1 2022–Q2 2026.

**Figure 2**. Forest plot of adjusted reporting odds ratios for pre-specified events, aflibercept versus faricimab and ranibizumab versus faricimab. Values below 1 indicate higher reporting with faricimab.

**Figure 3**. Time-to-onset distributions for the intraocular inflammation composite, by drug: cumulative incidence curves with Weibull shape parameters and median TTO.

**Figure 4**. Cross-database evidence classification heatmap: direction and significance of each signal across FAERS, JADER and EudraVigilance.

**Figure S1.** PT-level disproportionality signals for faricimab, aflibercept and ranibizumab versus the full FAERS database. Volcano plot; each point is one PT, x-axis log□ ROR, y-axis −log□□ FDR-adjusted P value. The 36 pre-specified non-AE PTs were excluded before analysis.

**Figure S2.** Adjusted versus unadjusted RORs at the SOC level. Side-by-side estimates for SOCs, faricimab as reference.

**Figure S3.** Adjusted versus unadjusted RORs at the PT level. Side-by-side estimates for PTs, faricimab as reference.

**Figure S4.** Systemic SOC-level head-to-head comparison, aflibercept versus faricimab.

**Figure S5.** Systemic SOC-level head-to-head comparison, ranibizumab versus faricimab.

**Figure S6.** Systemic SOC-level head-to-head comparison, ranibizumab versus aflibercept.

**Figure S7.** Cumulative time-to-onset curves, all adverse events.

**Figure S8.** Cumulative time-to-onset curves, arterial thromboembolic events.

**Figure S9.** Cumulative time-to-onset curves, death. The apparent leftward shift of the faricimab curve reflects its short post-marketing window (right-censoring), not a pharmacological difference.

**Figure S10.** Era-aligned first-three-years post-approval comparison. SOC-level RORs versus the contemporaneous full-database background for ranibizumab (2006–2009), aflibercept (2011–2014) and faricimab (2022–2025), each restricted to its first three post-approval years.

**Figure S11**. Masking sensitivity analysis, SOC level. log□ RORs computed against the masked background (all anti-VEGF reports removed) versus the unmasked full database; dashed line is the identity line. Spearman ρ = 0.984.

**Figure S12**. Masking sensitivity analysis, PT level. log□ RORs computed against the masked background (all anti-VEGF reports removed) versus the unmasked full database; dashed line is the identity line. Spearman ρ = 0.9996.

**Figure S13.** Cross-database SOC-level RORs, aflibercept versus faricimab. FAERS versus EudraVigilance aggregate-count estimates with 95% CIs.

**Figure S14.** Cross-database SOC-level RORs, ranibizumab versus faricimab. FAERS versus EudraVigilance aggregate-count estimates with 95% CIs.

## Notes

### Competing Interest Statement

The authors have declared no competing interest.

