## Supplementary figures and images for "A Distinct Delayed-Onset Intraocular Inflammation Phenotype of Faricimab versus Aflibercept and Ranibizumab: Triangulated Pharmacovigilance Evidence from Databases of US, Europe and Japan"

### Figure S1.pdf

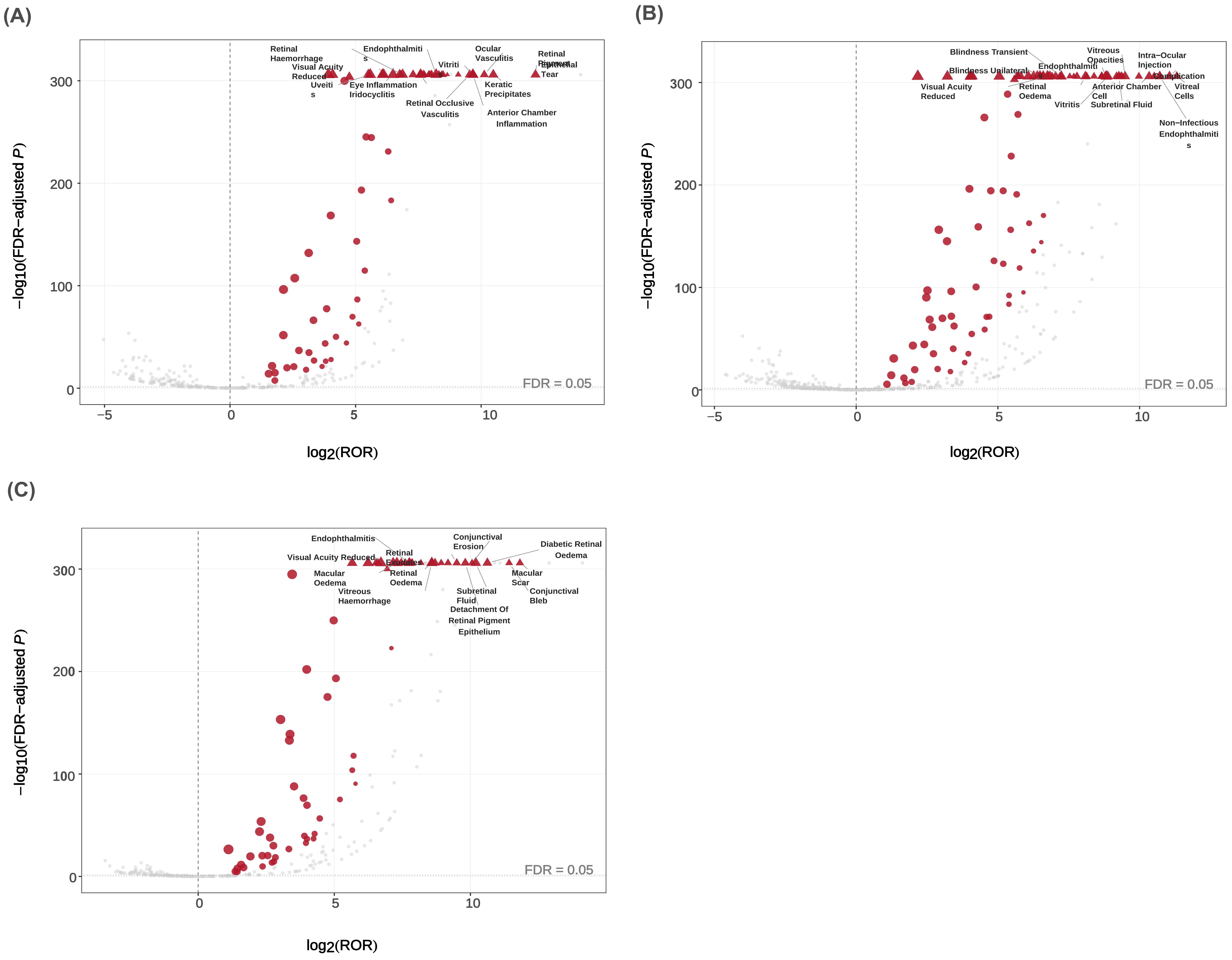

### Figure S2.pdf

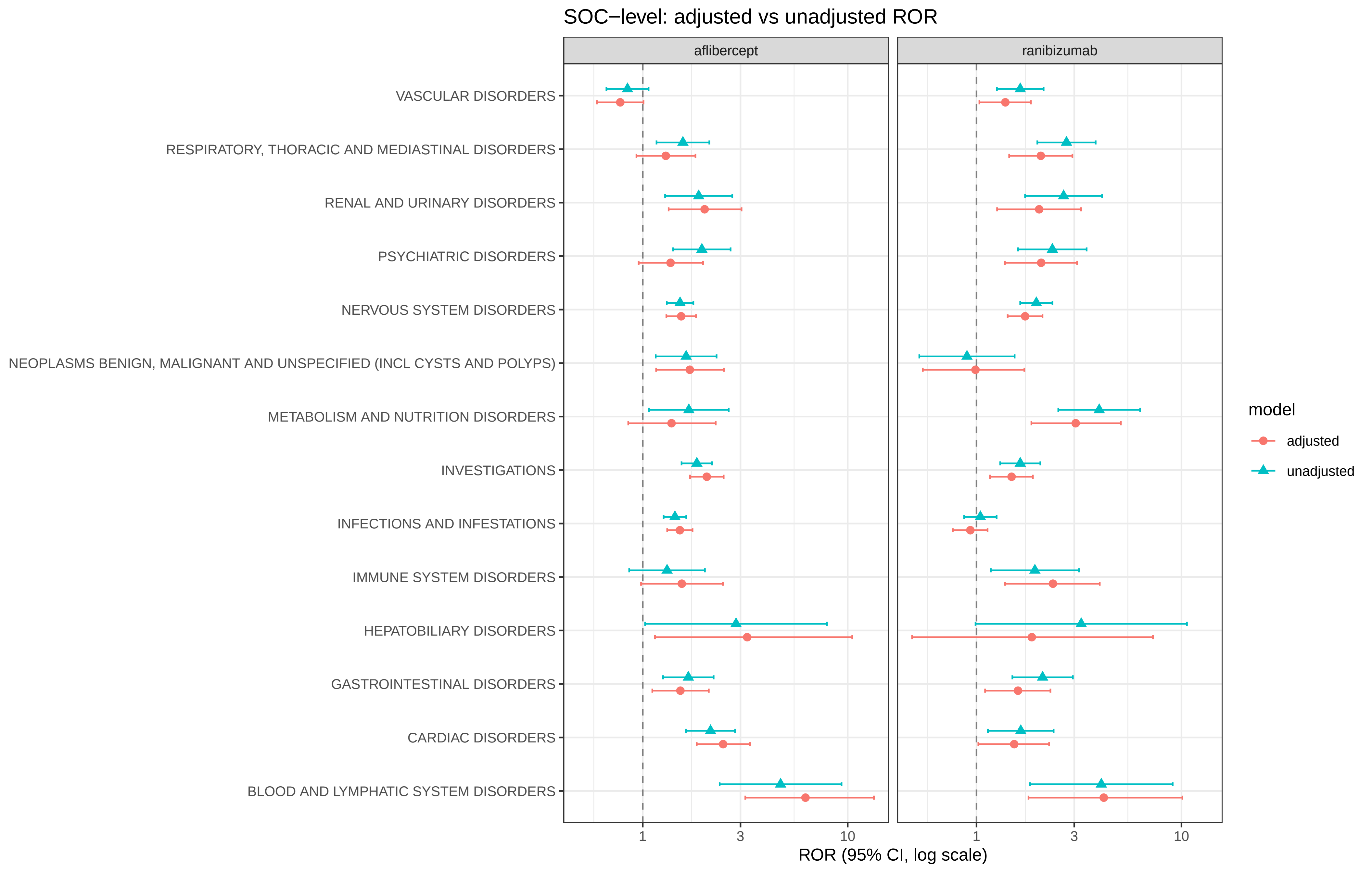

### Figure S3.pdf

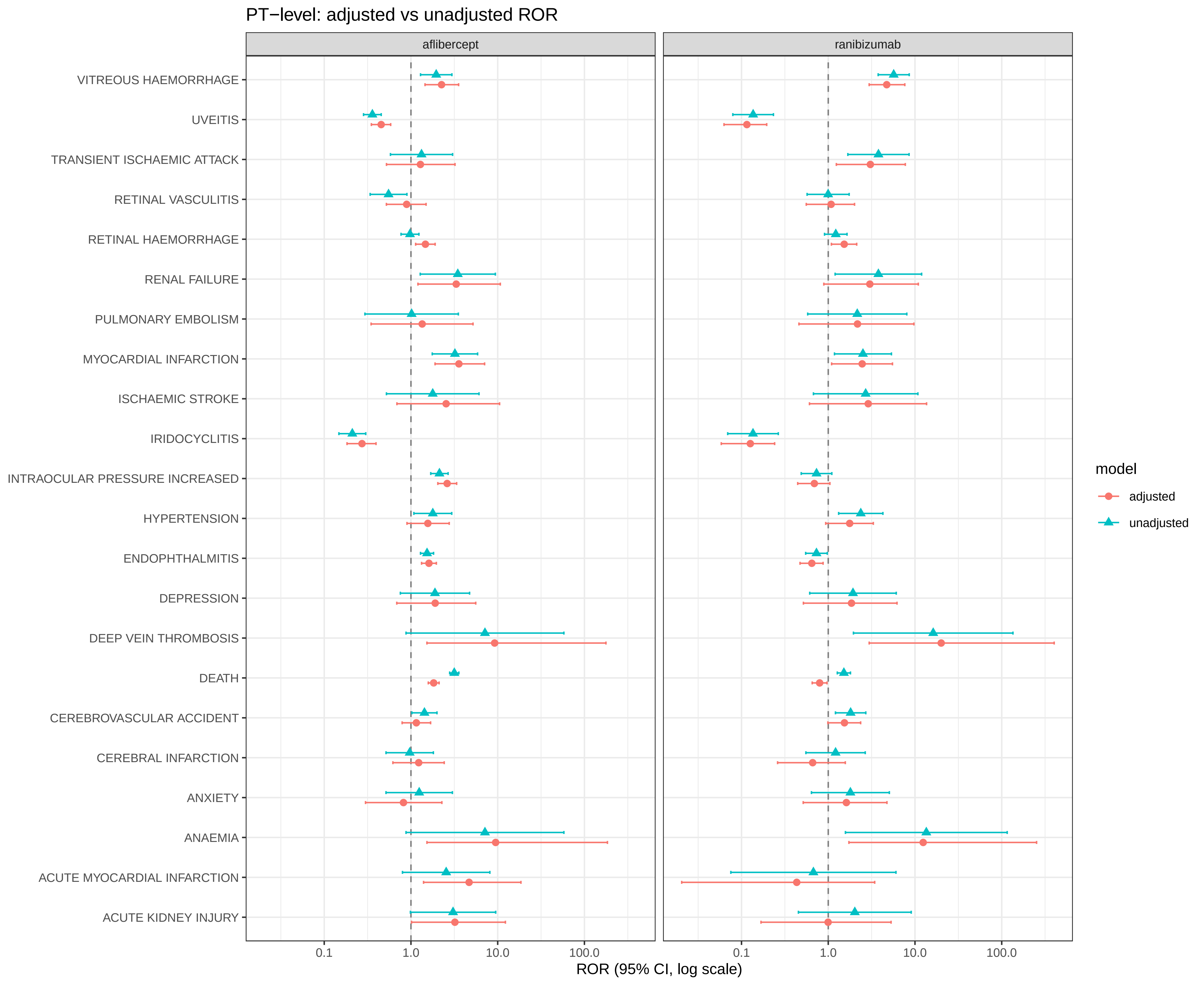

### Figure S4.pdf

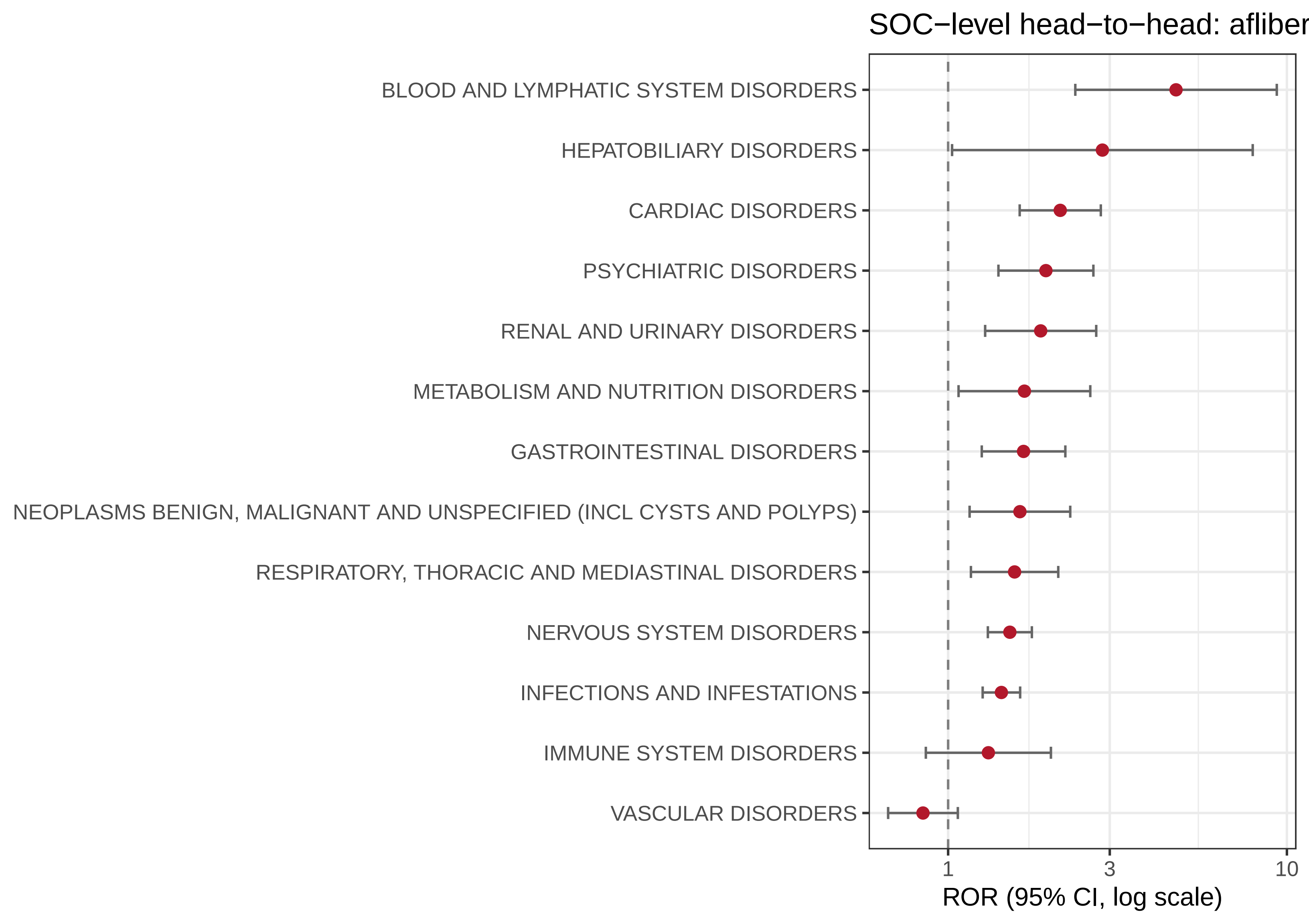

### Figure S5.pdf

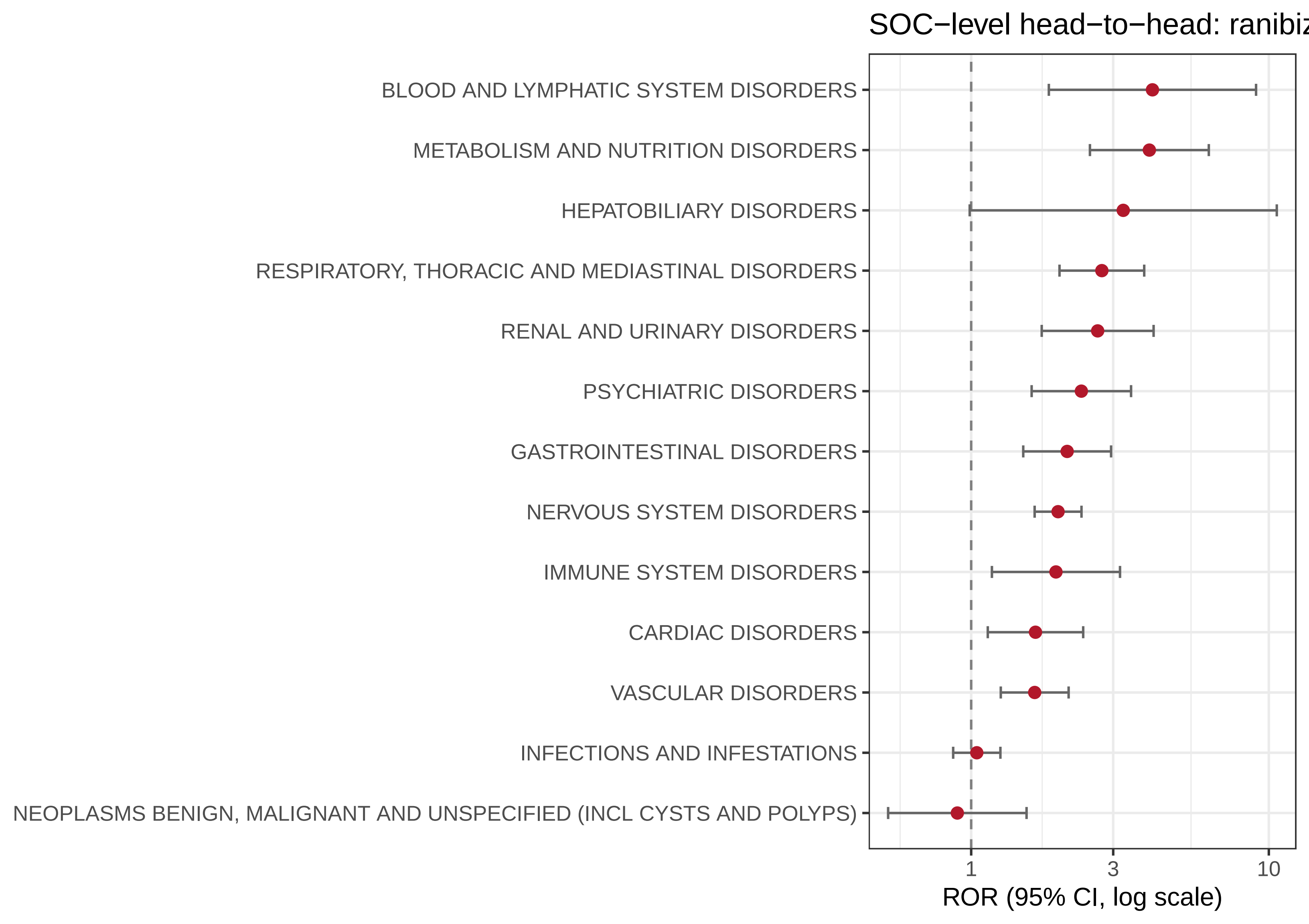

### Figure S6.pdf

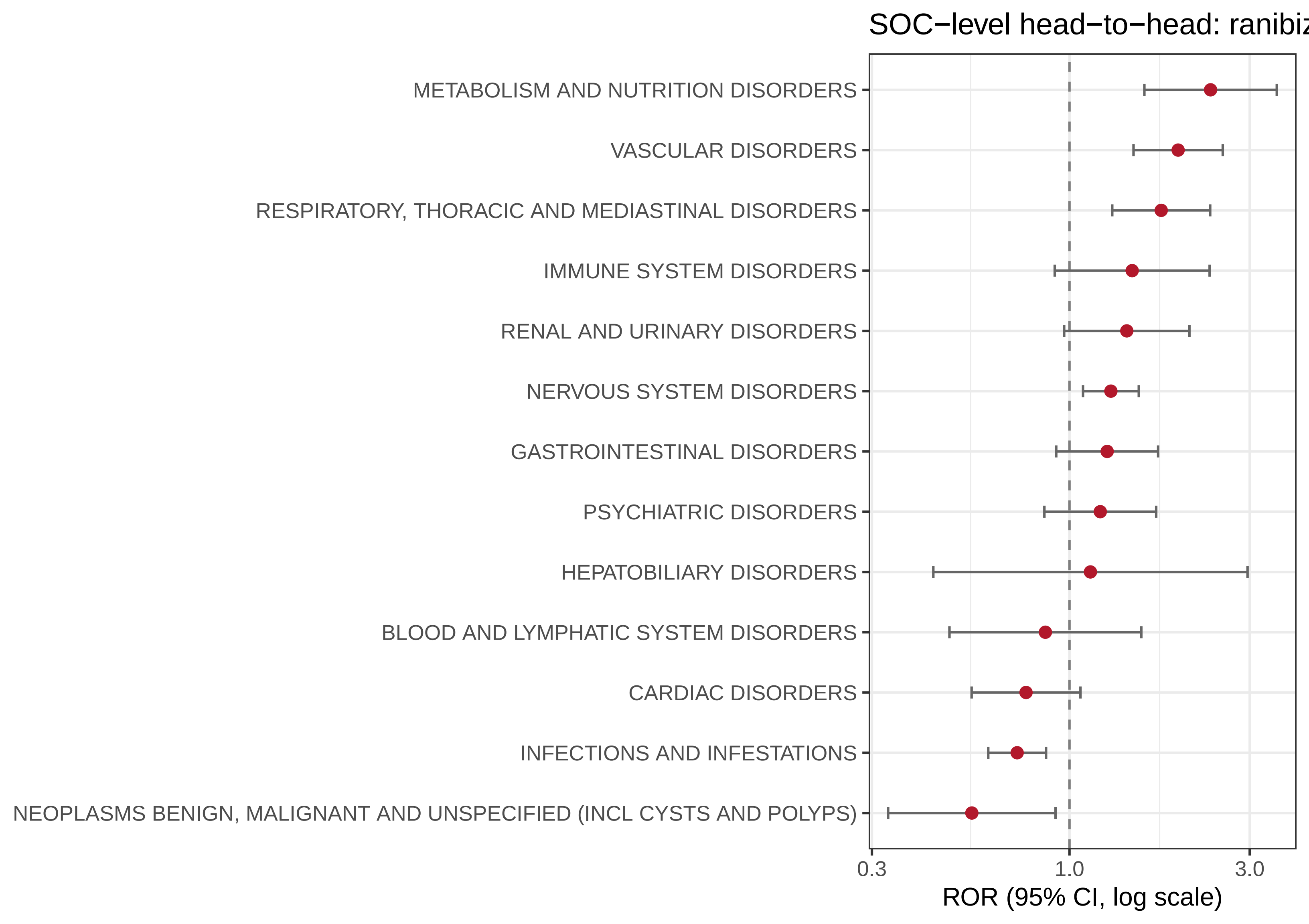

### Figure S7.pdf

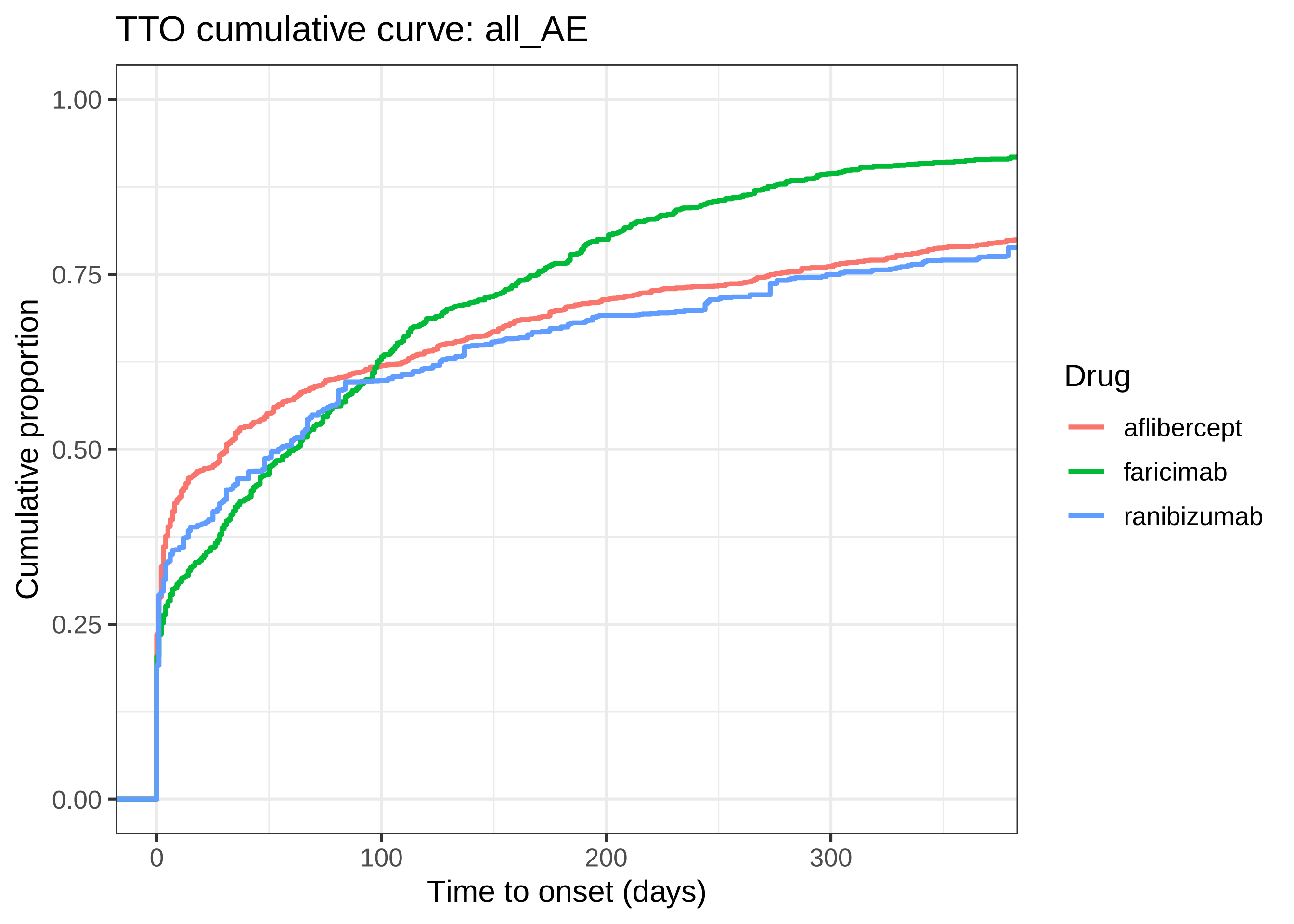

### Figure S8.pdf

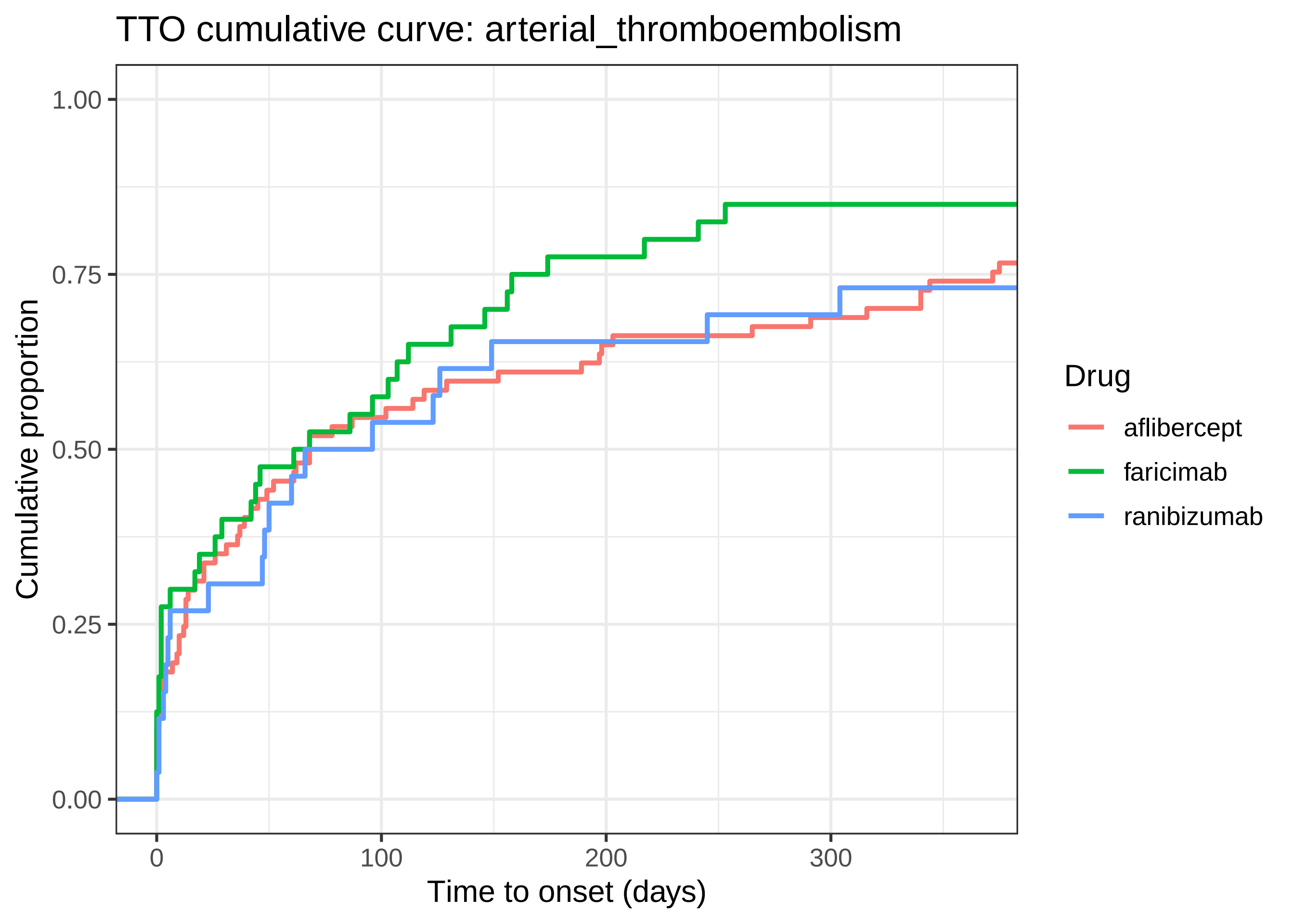

### Figure S9.pdf

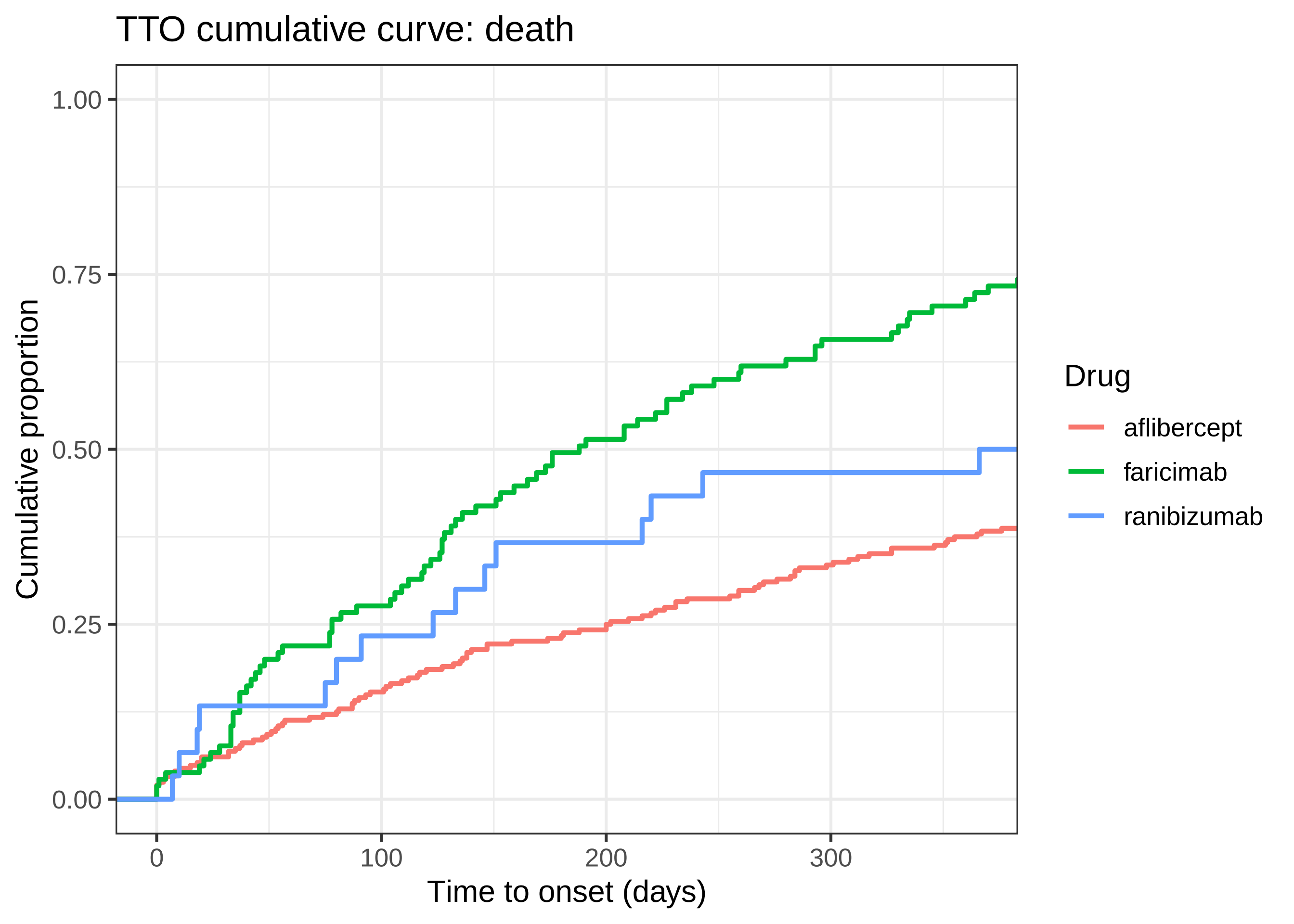

### Figure S10.pdf

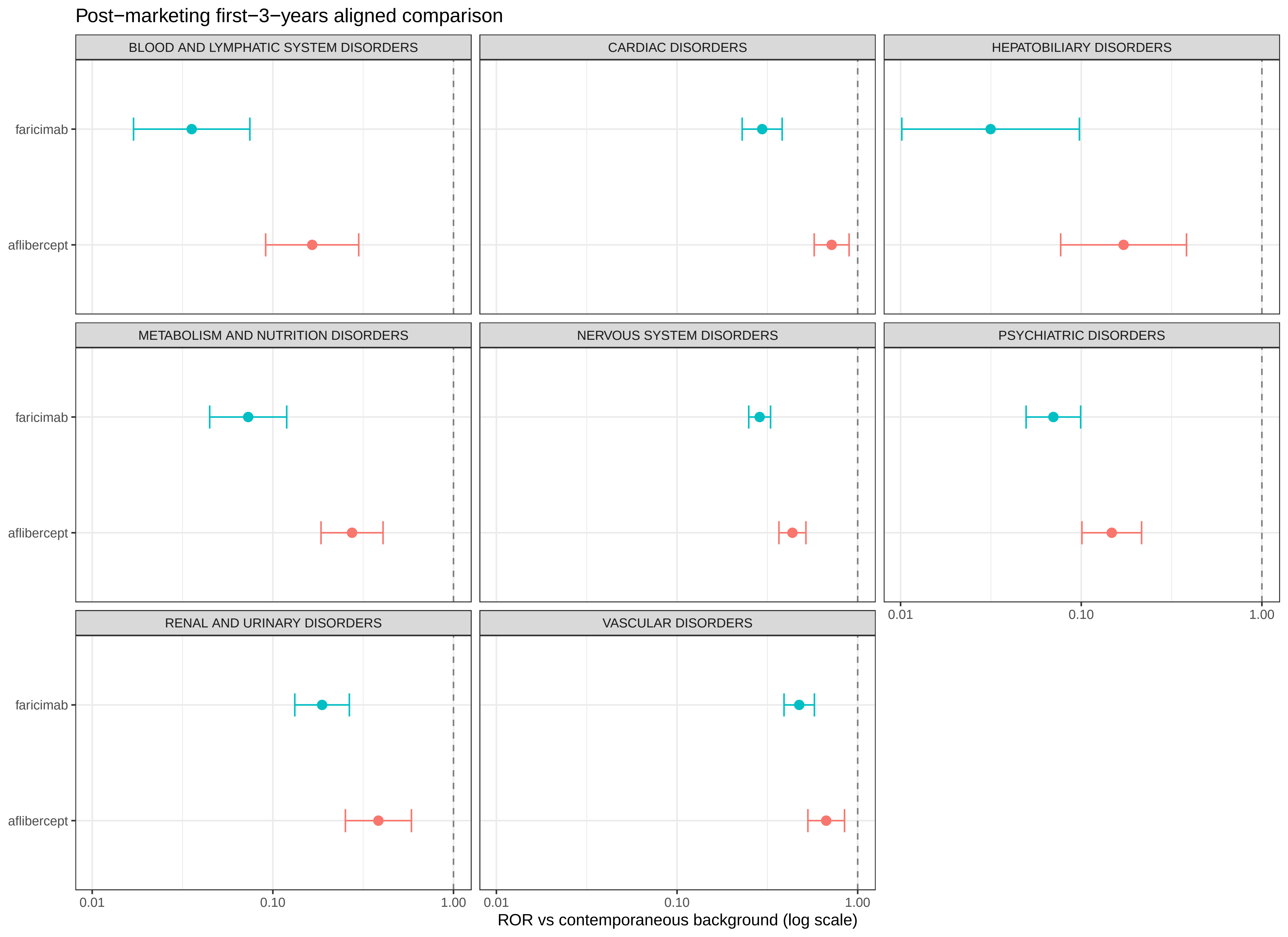

### Figure S11.pdf

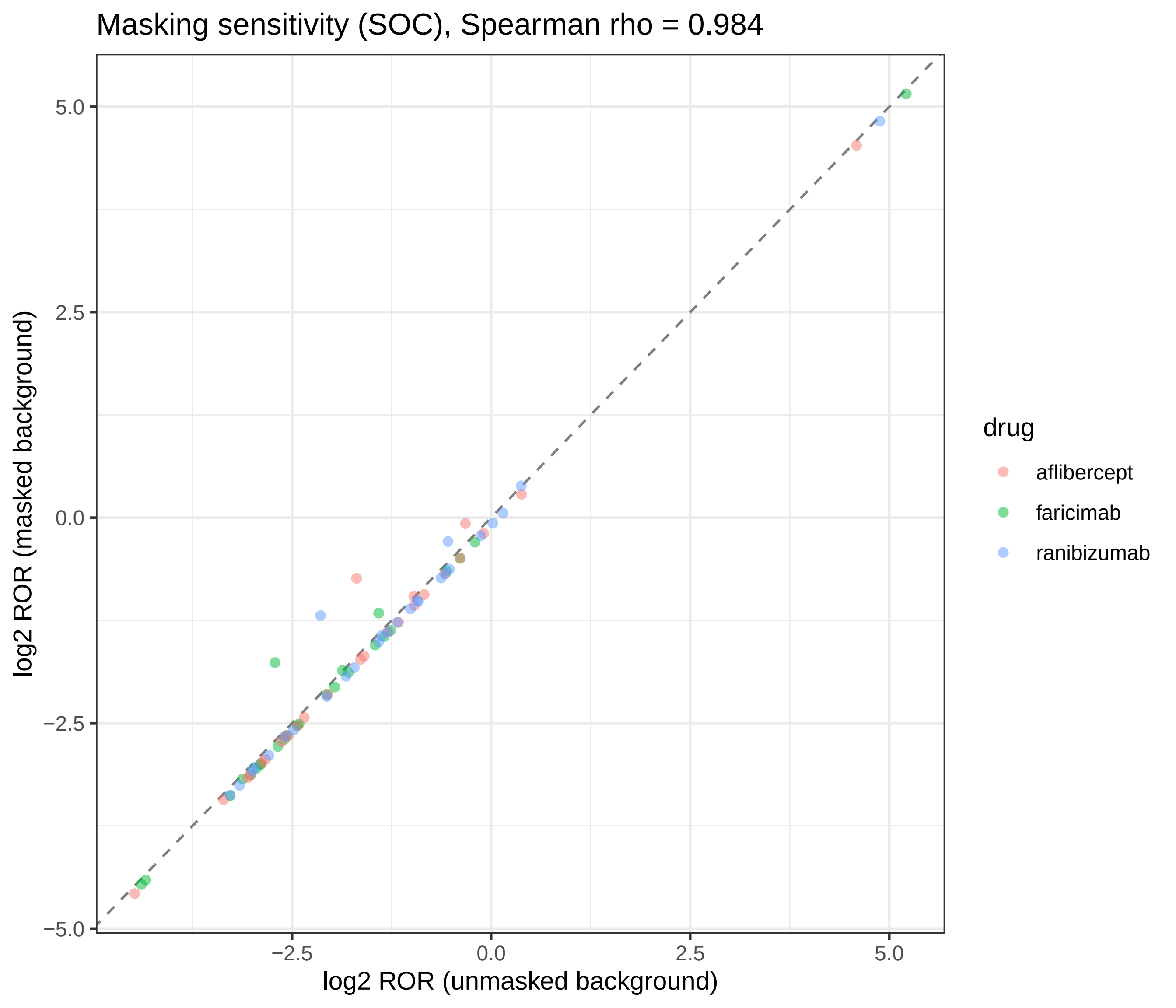

### Figure S12.pdf

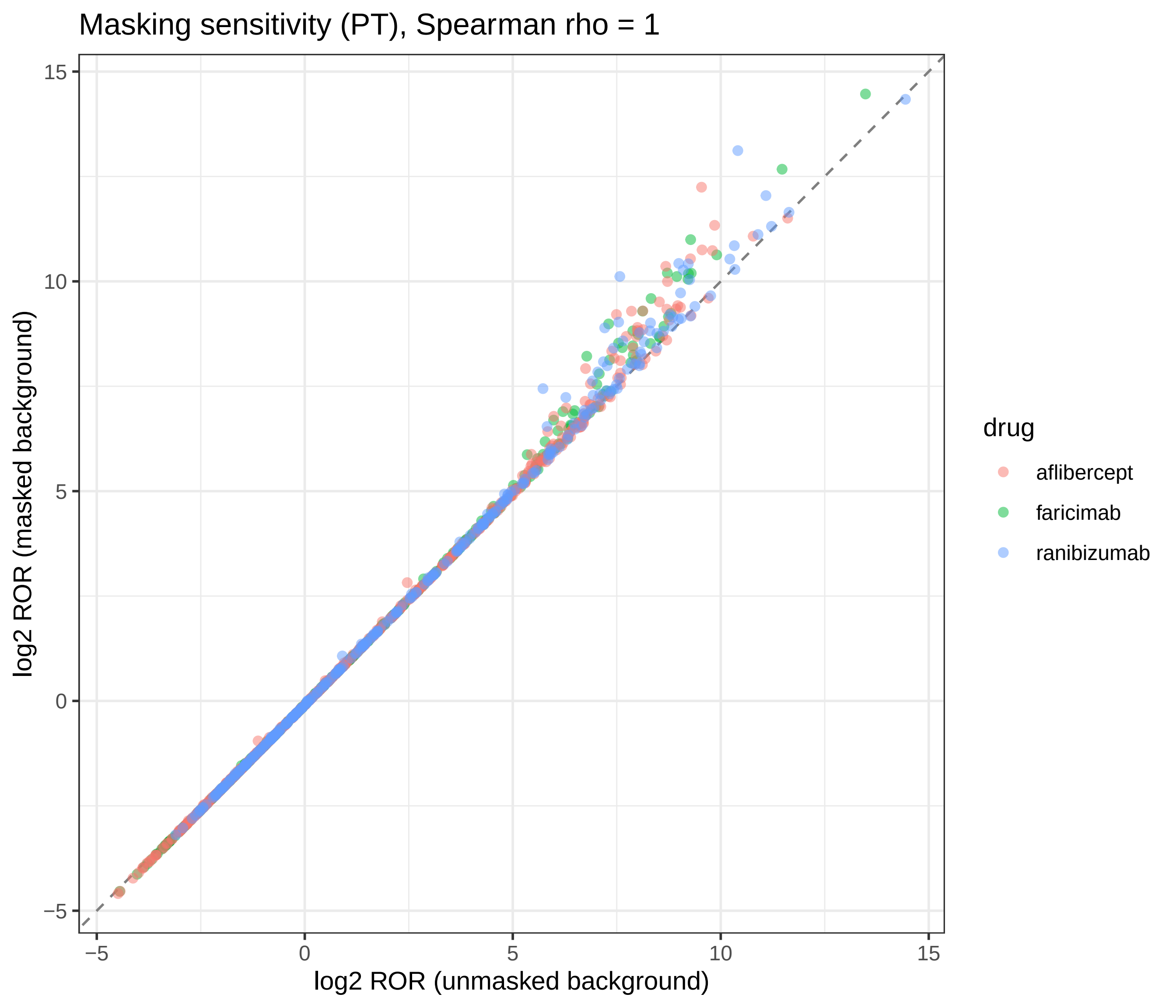

### Figure S13.pdf

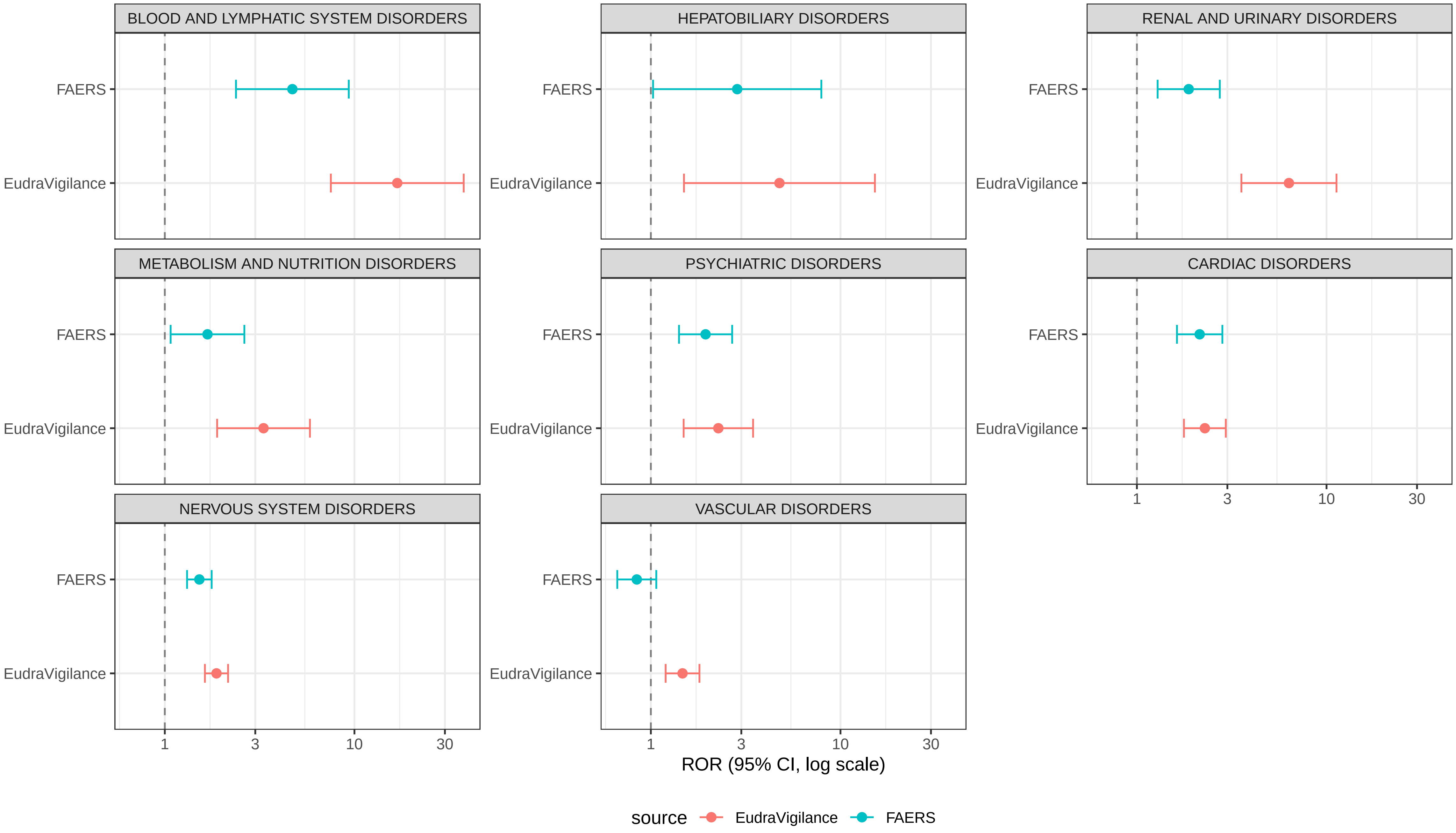

### Figure S14.pdf

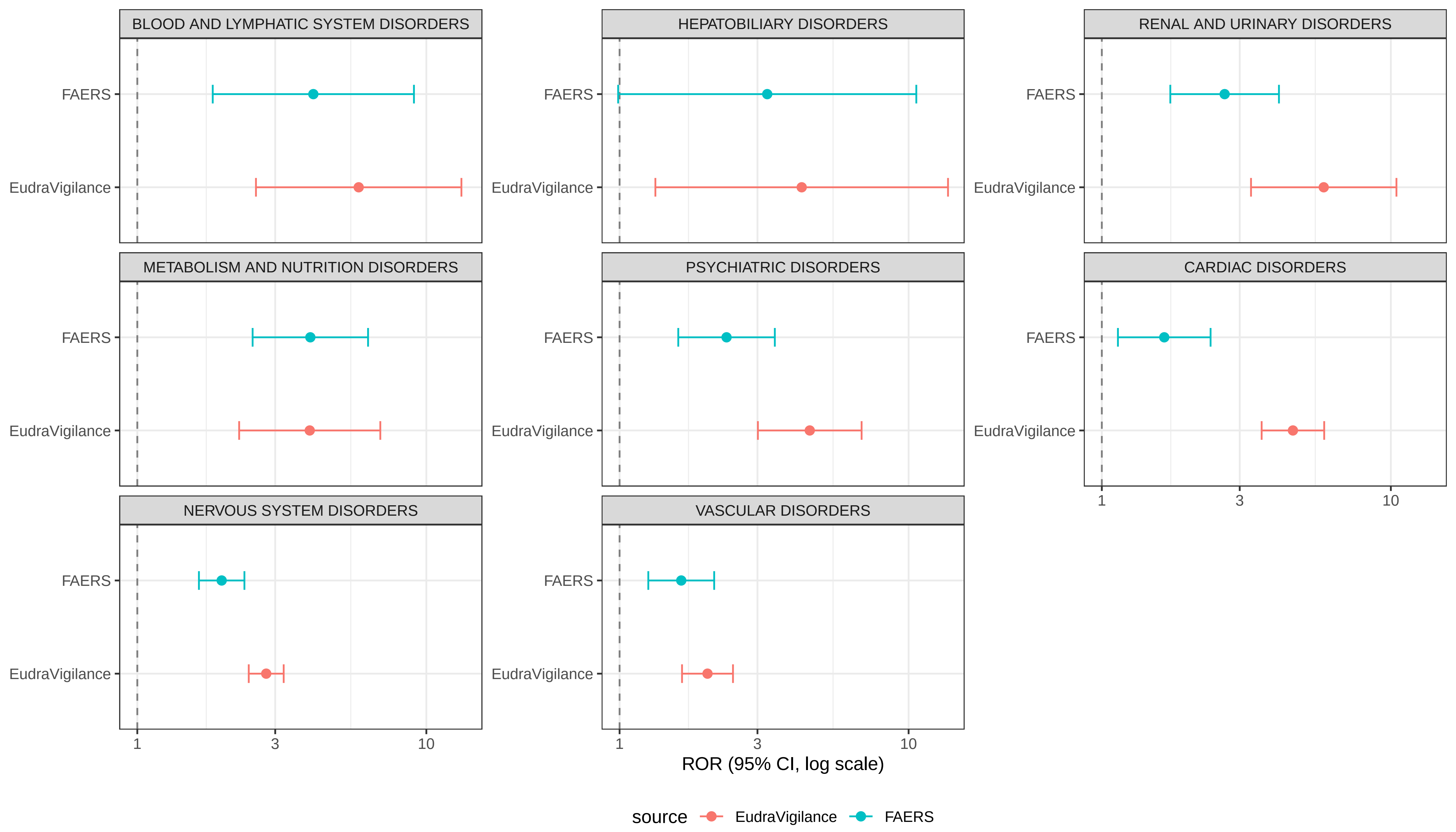
